# HEPATOCELLULAR CARCINOMA KINOME ATLAS AS AN EMPLOYABLE INSTRUMENTED PERSONALIZED MEDICINE

**DOI:** 10.1101/2025.07.04.25328828

**Authors:** Zachary A. Kipp, Evelyn A. Bates, Genesee J. Martinez, Wang-Hsin Lee, Sally N. Pauss, Terry D. Hinds

## Abstract

Liver cancer ranks high among cancer death rates and is resistant to most therapies. Studies of protein functionality are limited in patients with hepatocellular carcinoma (HCC). Here, we constructed an HCC kinome activity atlas and used it to develop personalized medicine technology applicable to profiling protein functionality. We used PamGene PamStation kinome technology that quantified protein kinase activity across hundreds of pathways for HCC tumor and non-tumor regions from both sexes. According to our bioinformatic analyses of kinases, ABL was the most active for men and women with HCC. Next, we employed three ABL inhibitors while running the PamStation, which revealed discernible alterations in ABL and other pathways. We deconvoluted over 500 kinase pathways and generated a personalized medicine (PerMed) score delineating activity levels; results were substantiated in five human hepatocyte cancer cell lines. Our work establishes an HCC kinome atlas and demonstrates PamStation’s potential for precision medicine applications.

## INTRODUCTION

Hepatocellular carcinoma (HCC) represents the most prevalent form of primary liver cancer, occurring in over 80% of cases ^1,2^. The overall five-year survival rate for individuals diagnosed with HCC is less than 20%, with this percentage declining to below 3% for advanced stages of the disease ^1,3^. Such a dismal prognosis is particularly concerning, especially given the global epidemic of hepatic dysfunction associated with obesity, referred to as metabolic dysfunction-associated steatotic liver disease (MASLD) ^4^, which significantly increases the risk of developing HCC ^5^. Obesity in youth elevates the risk of HCC in adulthood by 86–119% ^6^. The global prevalence of MASLD affects approximately 1.6 billion individuals ^7^, underscoring the alarming potential for a rise in future HCC cases. Unfortunately, HCC tumors are typically identified only at advanced stages, which are not suitable for liver transplantation. The standard treatment approach for advanced HCC is multimodal, necessitating a combination of pharmacological agents due to the high levels of resistance to therapy ^8^. An advantage of this study is that we quantified the HCC kinome atlas by profiling kinase activities across hundreds of pathways. We also developed personalized medicine technology that provides insights into the efficacy of therapeutic interventions for individual patients, including side effect profiles potentially enhancing their treatment success and quality of life.

The phosphorylation of proteins by kinases represents the most predominant form of posttranslational modification, comprising nearly one-third of all such alterations ^9^. The modification of phosphorylation events holds significant relevance to a myriad of diseases, including cancer, metabolic disorders, endocrine diseases, and numerous other conditions ^10–12^. Although various technologies are available to assess kinase pathways, they possess limitations and are not specifically tailored for comprehensive omics analyses of concurrent multi-pathway activities. Similarly, RNA sequencing (RNA-seq) pathway analysis fails to provide insights regarding the drivers of mRNA transcripts. However, an extensive and rigorous HCC investigation performed by The Cancer Genome Atlas Research Network analyzed 196 HCC samples for DNA methylation, RNA, miRNA, and proteomic expression and 363 HCC samples by whole-exome sequencing and DNA copy number analyses ^13^. This comprehensive work has been monumental in understanding gene expression in HCC. However, the functionality of the proteins was not tested. Therefore, methodologies that accurately determine the activity of kinases in real-time and the subsequent modifications in signaling pathways are crucial for advancing our understanding of how these pathways are altered in the context of disease.

Recent advancements in research technologies have focused on elucidating the activity of kinases, which are proteins accounted as master regulators of biological functions and physiological processes that also influence drug responses. Commonly employed methodologies for assessing kinase activity encompass immunoblotting, microarray technologies, and phospho-arrays. A state-of-the-art kinome microarray technology from PamGene has emerged called the PamStation. The difference between PamStation and traditional phospho-arrays is that the PamStation necessitates the inclusion of ATP in the cell or tissue lysate and acquires images during each step of the reaction, thereby allowing real-time simultaneous measurement of hundreds of substrates and kinase activities. The PamStation instrument utilizes PamChip microarray technology, which integrates peptide sequences incorporating kinase docking domains that facilitate substrate phosphorylation at phospho-tyrosine kinase (PTK) or serine/threonine kinase (STK) sites ^14^, exceeding three hundred forty (>340) pathways ^15^. A particularly promising application of the PamGene PamStation kinome technology resides in its potential use of analyzing these pathways within the domain of personalized medicine.

We published a study that used the PamGene kinome technology to investigate human subjects afflicted with cirrhosis, which is characterized as end-stage fibrosis. We conducted a comparative analysis of their kinase pathways against three rodent models of liver fibrosis. Our findings indicated that these subjects and rodent models exhibit analogous disease-related kinase signaling pathways, with the Discoidin Domain Receptor (DDR), a receptor tyrosine kinase (RTK) activated by collagen ^16^, and the insulin receptor (INSR) identified as the two most active PTK kinases ^10^. We delineated specific kinase activities within the PTK and STK families and assessed their implications in differentiating disease states from healthy controls. We recognized that this functional analysis and the associated data hold significant potential for enhancing predictions regarding treatment options and outcomes for HCC patients.

Therefore, we utilized the PamGene PamStation to measure the kinome in men and women with HCC. We profiled the real-time kinase activities from the phosphorylation of 340 STK and PTK substrates and performed an upstream kinome bioinformatic analysis comprising over 500 kinases. We hypothesized that by integrating STK and PTK data for each patient, we could evaluate the effectiveness and off-target side effects of various drugs against multiple diseases. To test this hypothesis, we incorporated drugs in the PamStation that target the primary pathway identified in our HCC patient sample kinome analysis. We coalesced the omics data to create an instrumented personalized medicine approach for a “Clinical Trial in a PamChip” that can be implemented per patient to enhance therapeutic responses, facilitate drug discovery, and address disease pathology.

## RESULTS

### Human HCC Tissue & Initial Kinase Substrate Sample Analysis

We employed liver biopsy specimens obtained from both cancerous (HCC tumor) and non-cancerous (normal adjacent tissue) regions from male and female patients diagnosed with HCC (**Figure 1A**). The characteristics of HCC patients, including body mass index (BMI), histological data, sex, race, and plasma levels of liver dysfunction biomarkers, specifically alanine transaminase (ALT) and aspartate aminotransferase (AST), as well as the HCC biomarker alpha-fetoprotein (AFP) ^5^, are detailed in **Supplemental Table 1**. Additionally, we validated the specimens for *GPC3* expression, a gene biomarker for HCC tumors ^17^. All specimens exhibited increased levels in the HCC tumors across both sexes, except one that had already been expressed highly in normal tissue (**Figure 1A**).

**Figure 1.**
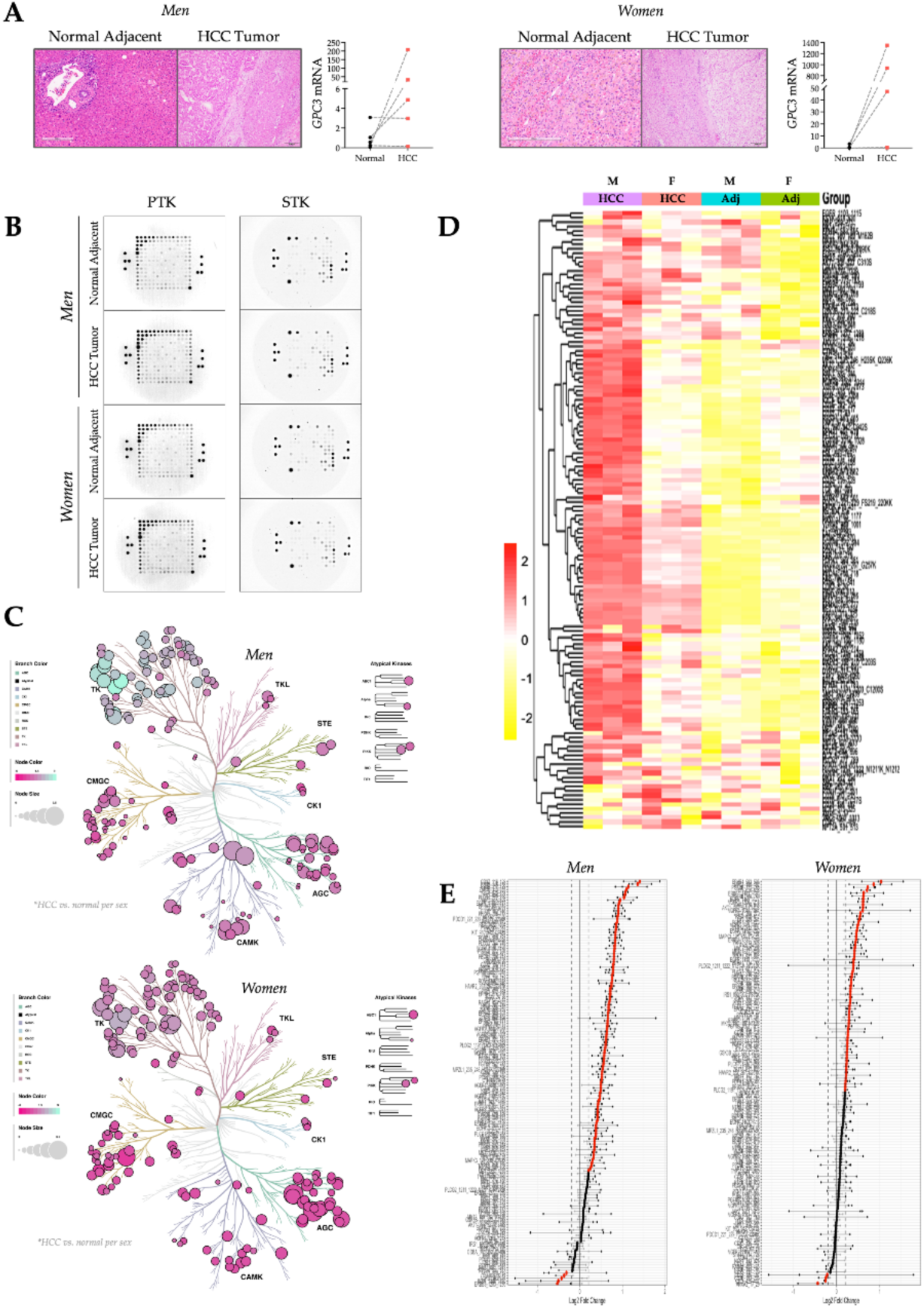
Human HCC tumors and initial kinase activity assessment. (A) H&E images of the men (left) and women (right) HCC tumor region and normal adjacent tissue (scale bar:) and *GPC3* mRNA expression [men, n=5, and women n=4 for tumor non-cancer (black circle) and (red square)]. (B) Representative image of phosphorylated PamChip after 94 cycles (PTK) or 124 cycles (STK) [men, n=5 and women, n=4 and samples were pooled and ran in triplicate for tumor or normal adjacent control]. (C) Phyla trees displaying the alterations of PTK and STK kinase activity in the HCC tumor vs normal adjacent for men (top) and women (bottom). (D) A heatmap showing the phosphorylation of 196 PTK substrates in the men and women HCC tumor and normal adjacent tissue. (E) A waterfall plot of the differentially phosphorylated PTK substrates in the men and women comparing HCC to normal adjacent tissue for each sex.

To determine the kinase activities in these samples, we utilized the PamGene PamStation PTK and STK PamChip microarray technology. We prepared liver biopsy sample lysates as we described for our human cirrhotic livers and other kinome analyses ^10,12,18–20^. Here, we used female and male HCC tumors alongside normal adjacent tissue as controls. We added the liver lysates to the kinome reagent mixture to identify kinase signaling differences, implementing patient proteins in the PamGene PamStation using the PTK and STK PamChip kinome technology. Illustrating the final phosphorylation level of the substrates in HCC compared to normal adjacent tissue, **Figure 1B** is a representative last image displaying microarray phosphorylation sites in the PTK (image #94) and STK (image #124) PamChips captured at the end of the run. We placed the calculated kinase data from the PTK and STK runs totaling 340 substrates into phylogenetic trees for visualization across sexes. Some overlaps were noted in the altered pathways for the kinase families TKL, STE, CK1, CAMK, and CMGC (**Figure 1C**). Combining the substrate data from the PTK PamChips into a heatmap reveals distinct phosphorylation levels between HCC and normal adjacent tissues for men and women (**Figure 1D**). The adjacent tissues from both sexes exhibited similar phosphorylation levels, with distinct changes in specific substrates. Overall, the substrate phosphorylation levels in female HCC samples were lower than in male HCC samples. Nevertheless, the TK substrates were differentially phosphorylated for certain pathways in men as opposed to women. **Figure 1E** depicts a waterfall plot of the PTK kinase substrates affected in HCC for men and women, which indicates the highest and lowest kinase substrate phosphorylation for both sexes.

### Upstream Kinase Assessments

We performed a deeper bioinformatic analysis to deconvolute the kinases that prompted substrate phosphorylation using the PamGene BioNavigator Upstream Kinase Analysis (UKA) software and many others described below. We represented the upstream kinase data derived BioNavigator UKA in waterfall plots for the PTK and STK PamChips (**Figures 2A & 2B**). The BioNavigator upstream kinase data from men and women with HCC indicate that PTK kinases were higher in HCC than in normal adjacent controls, which are represented by the line at zero, and in comparing the top ten kinases that exhibited the highest changes in the waterfall plots revealed that nine of these kinases were elevated in HCC of both sexes (**Figure 2A**). According to the BioNavigator UKA hierarchical rating, some PTK kinases in the top 10 have known HCC roles; the ABL kinase activity was the sixth highest in men and eighth in women (indicated by the blue arrow). ABL has a well-established role in causing HCC, and higher levels lead to worse prognosis and increased death rates ^21^.

**Figure 2.**
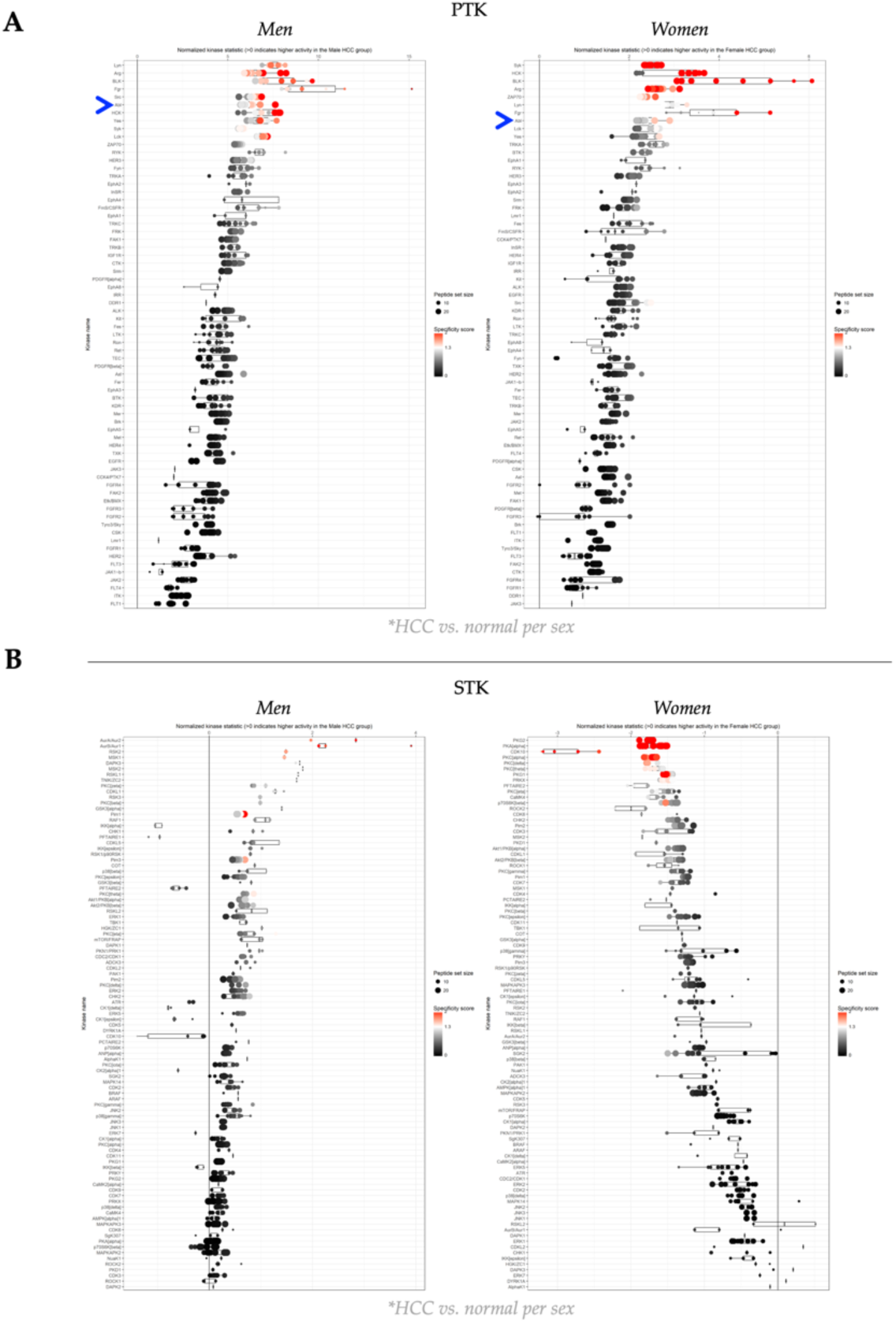
HCC tumors from men and women and upstream kinase assessments. (A) Waterfall plot of the PTK Upstream Kinase Analysis (UKA) in men and women comparing HCC to normal adjacent tissue for each sex. Blue arrow denotes the location of ABL. (B) Waterfall plot of the STK Upstream Kinase Analysis (UKA) in men and women comparing HCC to normal adjacent tissue for each sex.

The STK waterfall plot analysis suggested a dichotomy between men and women with HCC, as men had higher STK activity and women reduced (**Figure 2B**). Several members of the Protein Kinase C (PKC) family were elevated in males and reduced in females; some families exhibited similar regulatory actions, such as decreased PFTAIRE1 and PFTAIRE2 kinase activities in both sexes. A surprising finding was that AKT1 and AKT2 in men showed increased kinase activity; in women, they were reduced. However, the PTK data for the insulin receptor (INSR) indicated higher kinase activity in both men and women (**Figure 2A**). The changes in mRNA levels between cancerous and non-cancerous samples were mostly similar among both sexes (**Supplemental Figure 1**), except for GSK3, which was higher in women but not men.

Validation of the expression level of some of the most changed kinases showed that INSR, SYK, AKT1, AKT2, and ERK1 (*MAPK1*) had similar mRNA changes between normal and HCC tumor samples in both sexes, indicating that it is indeed the protein functionality that was considerably changed.

### Kinase Hierarchical Computations

To further interrogate the PTK pathways, we analyzed the kinase activity data by Z-score for each sex. The Z-score waterfall plot data analysis indicates that ABL was the most active PTK kinase in HCC for both sexes (**Figure 3A**). The KRSA upstream analysis also indicated ABL as men’s most active kinase and women’s second most active after INSR (**Supplementary Table 2**). Using the Z-score waterfall plot, INSR was the second most active kinase for women and the eighth for men (**Figure 3A**). On the Z-score waterfall plot, SYK ranked fourth for men and fifth among women; the KRSA analysis showed SYK ranked fourth among men and third among women. **Figure 3B** depicts a waterfall plot of the ABL substrates affected in HCC for both men and women.

**Figure 3.**
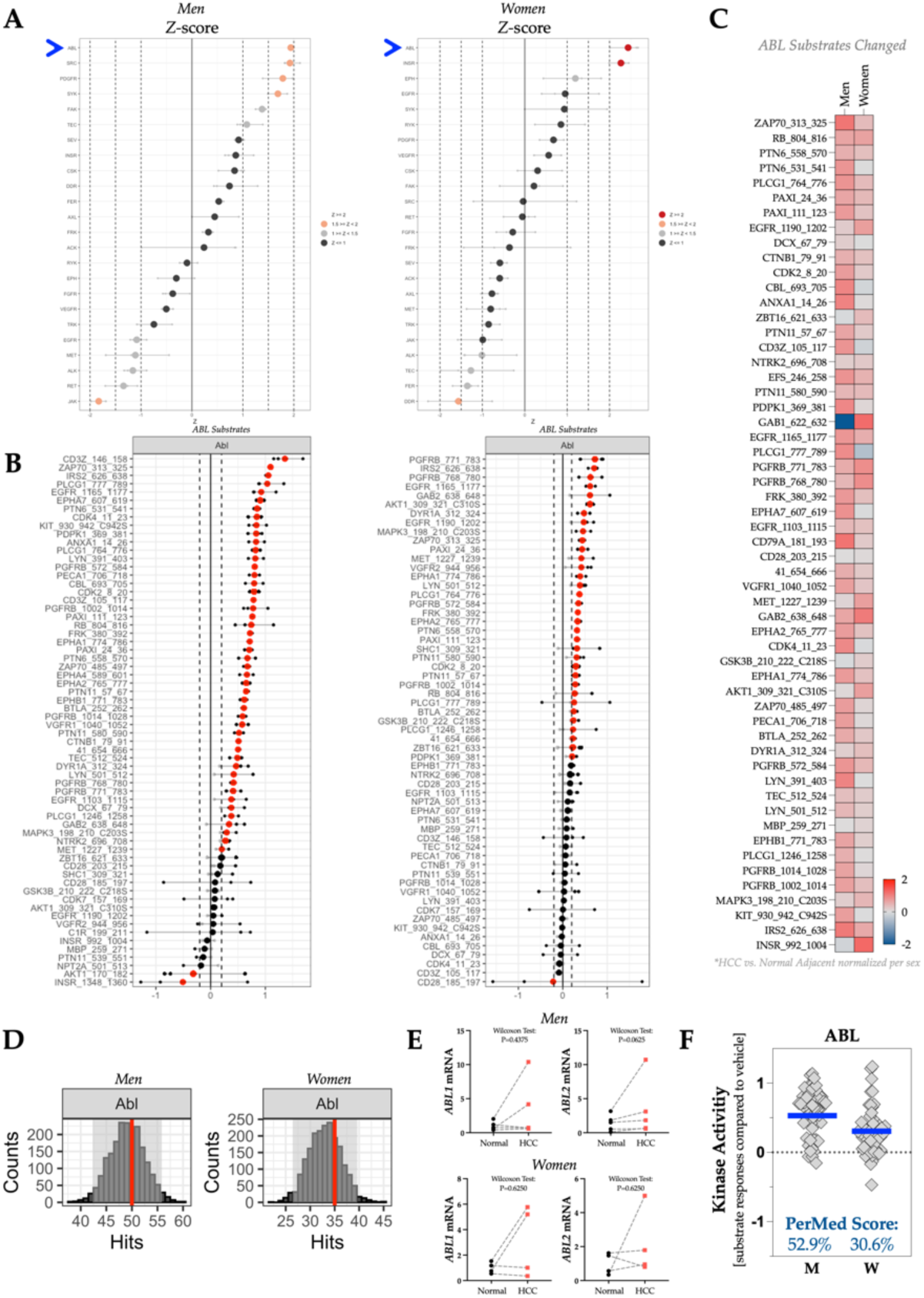
Kinase hierarchical computations of HCC tumor for target pathway. (A) Z-score waterfall plots of the upstream PTK kinases in the HCC tumor vs normal adjacent tissue. The blue arrow denotes the location of ABL. (B) Waterfall plot of the ABL substrates in the HCC tumor compared to normal adjacent tissue in men (left) and women (right). (C) Heatmap of the phosphorylation level of ABL substrates. (D) Peacock plots of ABL in HCC tumor vs normal adjacent. (E) RT-PCR of ABL1 and ABL2. (F) The kinase activity of ABL and the PerMed Score denoting the percent change in kinase activity in the HCC tumor compared to normal adjacent. The blue line denotes the average signal intensity of each substrate compared to the control.

A heatmap comparison of ABL substrate phosphorylation levels between men and women indicates similar phosphorylation for most ABL substrates, with only a few not aligning (**Figure 3C**). To investigate ABL further, we performed random sampling analyses and quantified gene expression changes between the two sexes. In **Figure 3D**, the peacock plots, measuring thousands of iterations of random peptide selection as we described previously ^10^, signify that the number of differentially phosphorylated peptide substrates observed, shown by the red line, has high confidence. The *ABL1* and *ABL2* mRNA levels in HCC compared to normal adjacent tissue did not significantly change in men or women per the Wilcoxon test for analyzing paired data (**Figure 3E**). The personalized medicine score (PerMed Score) is calculated by comparing the control sample to the percentage of change in the experimental group. It reflects changes in the activity of specific kinases, which could be in a disease state, drug treatment, or other investigations. The PerMed Score for ABL kinase activity is derived from all ABL substrates combined for each sex, normalized to their controls (zero line), with the mean as a blue bar. The ABL PerMed Score in men was 52.9% and 30.6% for women (**Figure 3F**), indicating that ABL kinase activity in both sexes was substantially elevated in HCC.

### Drug Efficacy Measures via Kinases

The comprehensive kinome analysis, which included PTK and STK data evaluations, revealed that women and men diagnosed with HCC exhibited hyperactivity of ABL compared to normal adjacent tissue. Based on these findings, we employed the same human liver lysates from both sexes to investigate the efficacy of inhibiting ABL activity through specific antagonists utilizing the PamGene PamStation instrument during a subsequent PamChip run with the same samples. The PTK and STK PamChip analyses were repeated while modifying the reagent mixture to incorporate three ABL inhibitors: imatinib, rebastinib, or olverembatinib. The HCC tumor protein lysates from both sexes were pooled to generate a kinome atlas of drug responses (**Figure 4**) and processed individually for each patient sample (**Supplemental Figures 2 & 3**).

**Figure 4.**
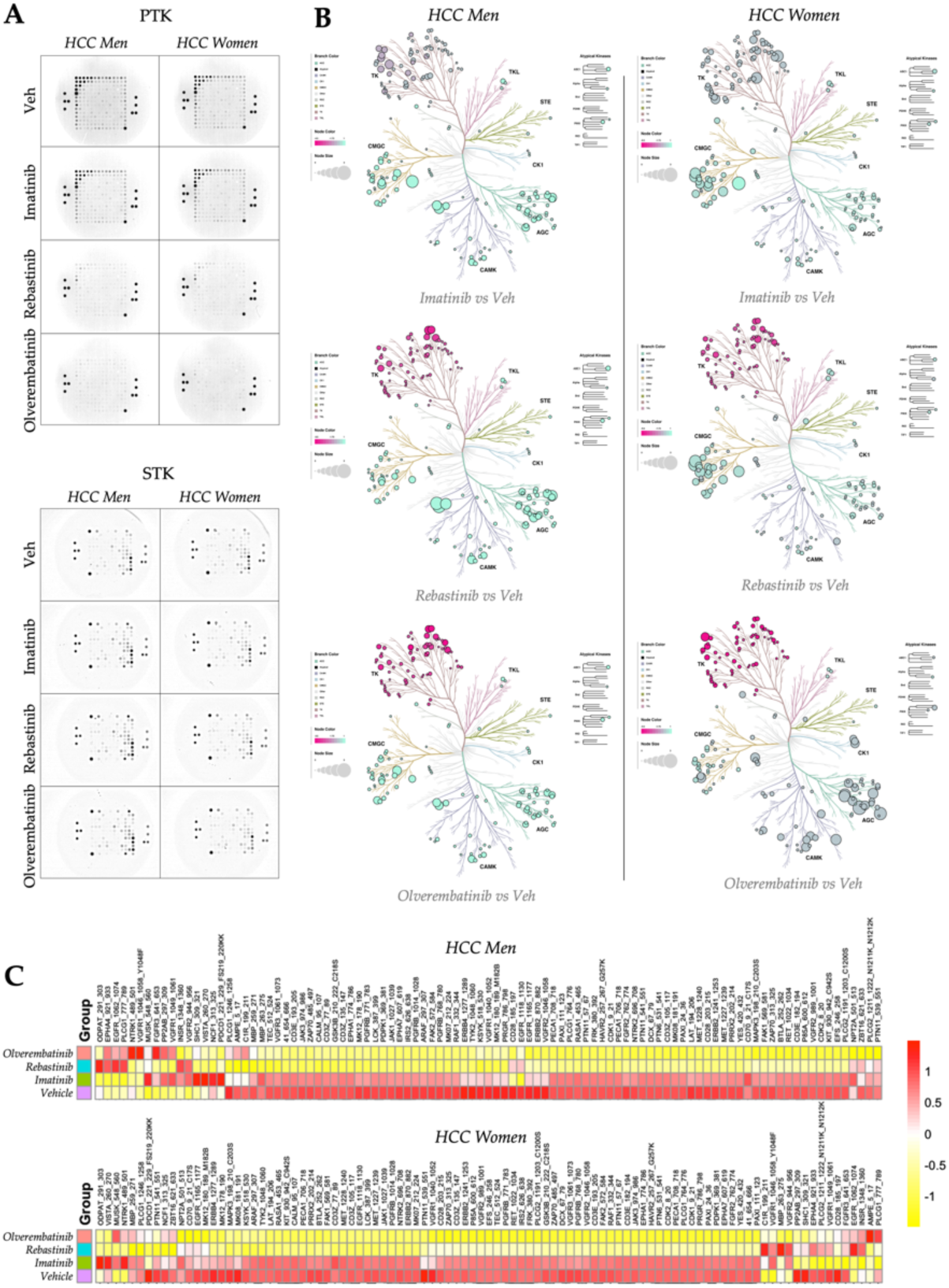
Drug efficacy quantification via kinases and ABL inhibitor signaling. (A) Images of phosphorylated PTK (top) and STK (bottom) PamChip during the final cycle. (B) Phyla trees displaying the alterations of PTK and STK kinase activity in the HCC tumor with Imatinib (top), Rebastinib (middle), or Olverembatinib (bottom) compared to vehicle in men (left) and women (right). (C) Heatmaps showing the phosphorylation of 196 PTK substrates in men (top) and women (bottom) HCC tumor samples with vehicle, Imatinib, Rebastinib, or Olverembatinib.

The human HCC tumor protein lysates previously prepared were snap frozen and stored at -80 C and reused for the individual kinome analysis once the primary target was identified, which was ABL in men and women. Hence, we used ABL inhibitors to conduct a “Clinical Trial in a PamChip.” A similar PamGene PamStation PTK and STK kinome running protocol was employed with two modifications. The same amount of protein lysate mentioned above was used, but the ATP concentration was reduced to ensure it would not outcompete the inhibitors used. The initial protein lysates were added with the same mass as before to a reagent mixture containing imatinib (1 µM final concentration), rebastinib (100 nM final concentration), olverembatinib (100 nM final concentration), or vehicle (DMSO) and incubated for 30 minutes at room temperature while gently rocking before adding ATP and a fluorescently labeled antibody. The final concentrations of the ABL antagonist compounds were determined by their inhibitory concentration 50 (IC50) values described on SelleckChem.com. A consistent final concentration of DMSO (2%) was applied across all treatments. The operational conditions for the PamGene PamStation remained aligned with those utilized in the initial run. The resultant raw images were exported for quantification and subsequent analysis. The microarray phosphorylation pattern observed in the PTK PamChip exhibited a slight decrease with imatinib and a substantial reduction when treated with rebastinib or olverembatinib across both sexes (**Figure 4A**). The phosphorylation of the STK PamChip displayed only marginal changes upon treatment with all three compounds. The integration of kinase activity data from the PamChips into phylogenetic trees for visualization of 340 substrates indicates that imatinib exerted modest effects on the TK family and had a similar influence on the STK kinases as rebastinib and olverembatinib (**Figure 4B**). In conjunction with the ABL inhibitors, a heatmap illustrating the PTK substrate data revealed vastly reduced phosphorylation levels with rebastinib or olverembatinib. At the same time, a slight decrease was observed with imatinib across both sexes (**Figure 4C**). We assessed kinase activity data and represented it in waterfall plots for the PTK and STK datasets specified in the comprehensive kinome analysis. Utilizing the BioNavigator UKA software in male and female subjects with HCC shows reduced pathways compared to normal adjacent controls. **Figures 5A and 5B** depict the specificity of the ABL inhibitors (noted by a blue arrow in PTK) using the hierarchal rating system from the BioNavigator UKA software.

**Figure 5.**
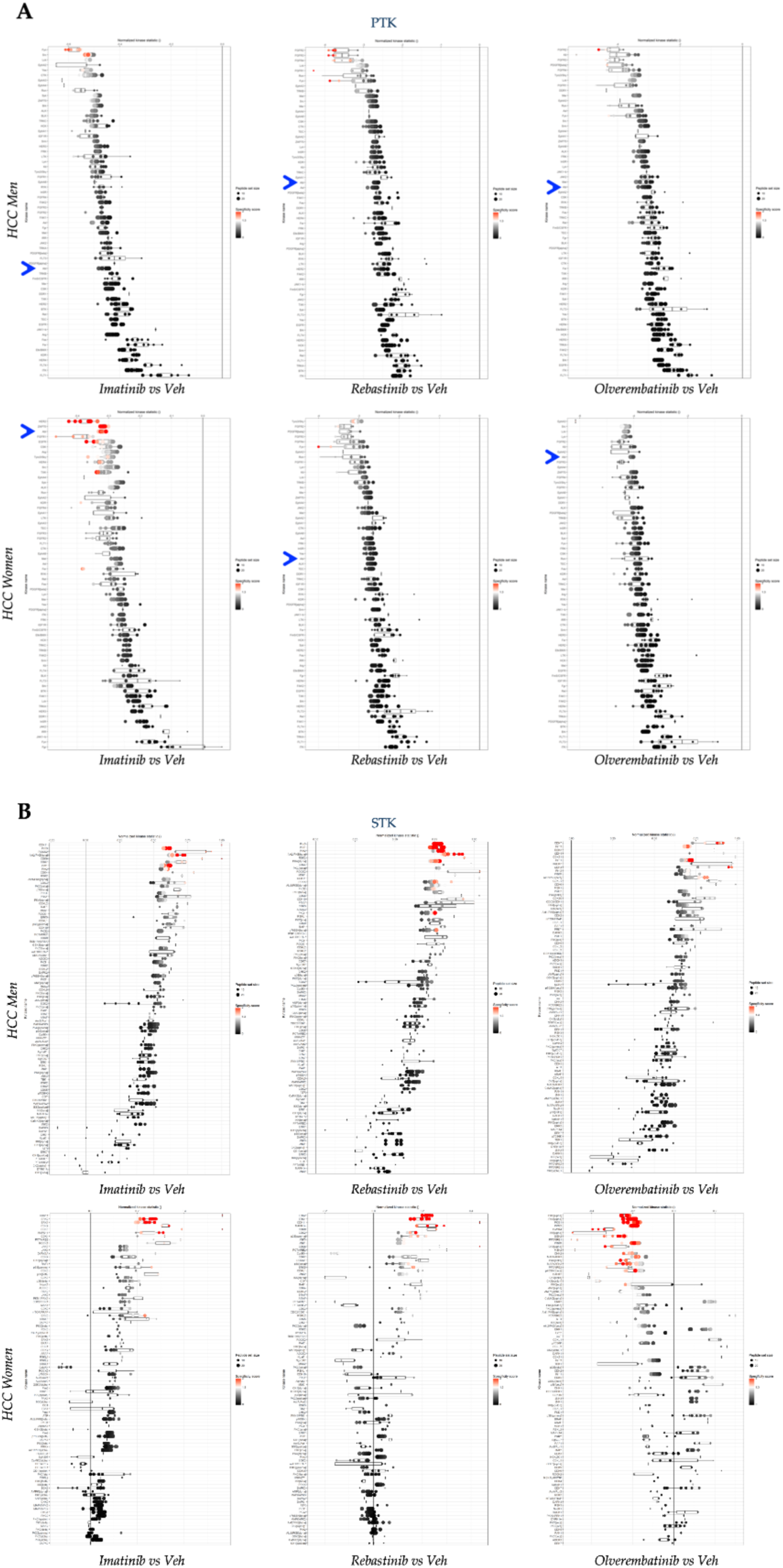
Specificity of drug actions on kinases. (A) Waterfall plot of the PTK Upstream Kinase Analysis (UKA) in men and women comparing Imatinib, Rebastinib, and Olverembatinib to vehicle for each sex. Blue arrow denotes the location of ABL. (B) Waterfall plot of the STK Upstream Kinase Analysis (UKA) in men and women comparing Imatinib, Rebastinib, and Olverembatinib to vehicle for each sex.

The activity of the ABL and several PTK kinases was diminished; however, the STK kinases exhibited a dichotomous response. The activity of multiple STK kinases was elevated in the presence of all three ABL inhibitors in men. The results varied in women, however, with imatinib enhancing the activity of certain STK kinases while diminishing that of others. Rebastinib also increased the activity of many STK kinases to a lesser extent than that observed with imatinib.

Conversely, Olverembatinib predominantly demonstrated a pattern of decreased STK kinase activities, with fewer instances of elevation. These findings suggest that the specificity of each compound varies, which was detectable using the employed methodology. Notably, pronounced sex differences were observed with all ABL antagonists. These differences will likely contribute to varying patient outcomes, possibly attributed to off-target side effects.

### Specific Drug Actions on Kinases

The waterfall plots of the ABL targets show substrates affected by the three ABL antagonist treatments, and the Z-score waterfall plot signifies that ABL was one of the most significantly targeted kinases in the HCC kinase populations for both sexes (**Figure 6A**). A heatmap comparison of ABL substrates phosphorylated in men and women shows comparable phosphorylation for nearly all ABL substrates, with imatinib having lesser actions (**Figure 6B**). **Figure 6C** demonstrates that ABL targets changed considerably with the ABL inhibitors in both men and women. Imatinib had the least effect on inhibiting ABL’s activity in male and female HCC samples (PerMed Score: -45.0% for men and -30.8% for women) compared to vehicle. However, rebastinib and olverembatinib significantly reduced ABL kinase activity, with PerMed Scores of -261.6% and -297.2% for men and -275.2% and - 401.4% for women. The antagonists suppressed the kinase activities of ARG/ABL2 compared to those of ABL activity levels. An interesting observation is that imatinib had a measurably lesser inhibitory action against proteins from HCC samples from women. There have been sexual differences reported for imatinib responsiveness in leukemia cancer patients ^22^, which might indicate that these molecular differences detected in our study may be related. Women have been shown to have significantly more relapse to imatinib therapy ^23^. Our analysis here may serve as an early detection tool for identifying these changes, especially if numerous other pathway changes are included as a hallmark of resistance.

**Figure 6.**
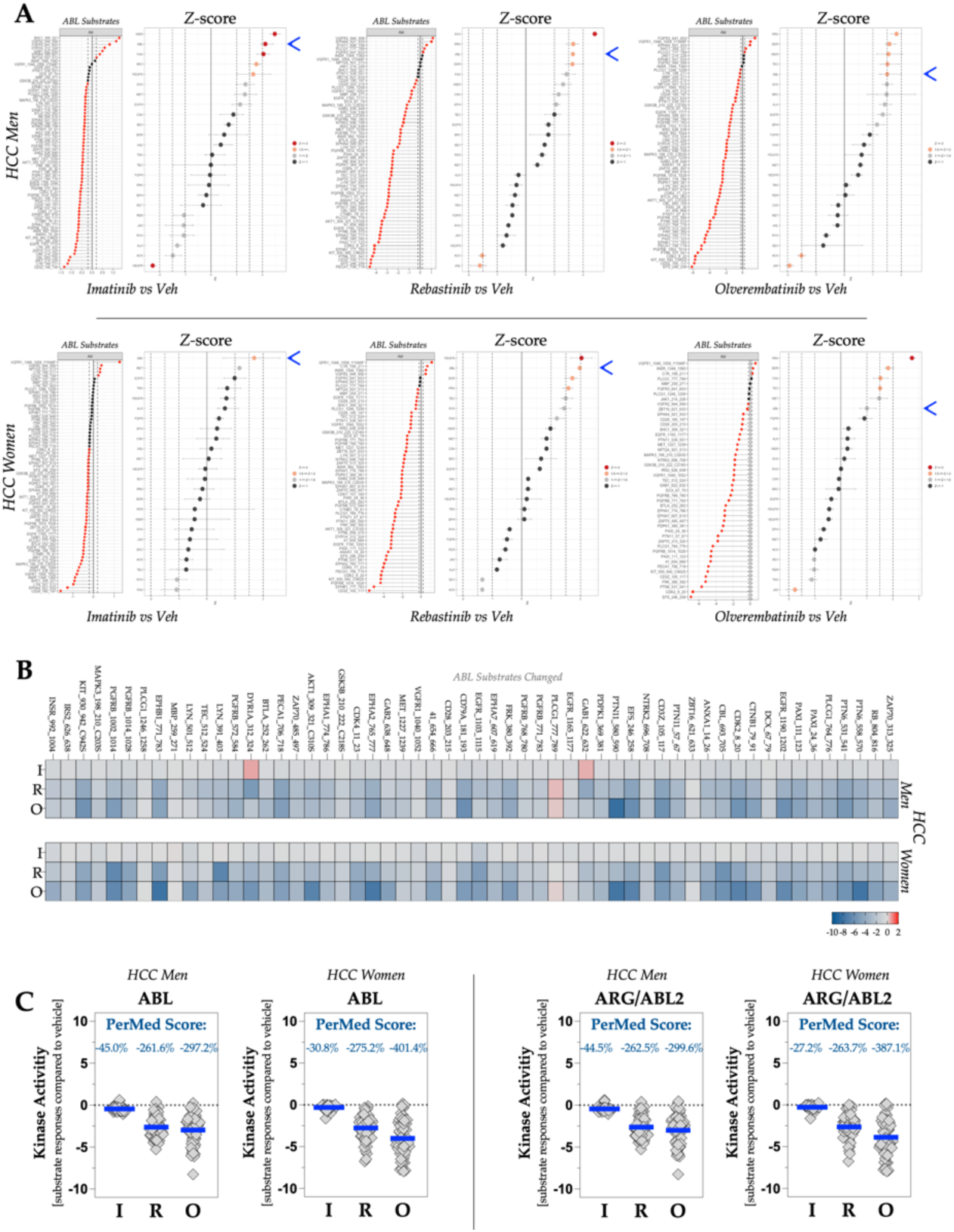
ABL antagonists and personalized medicine score analysis. (A) Waterfall plot of ABL substrates and Z-score plots of men (top) and women (bottom) HCC tumor with Imatinib (left), Rebastinib (middle), or Olverembatinib (right) compared to vehicle. Blue arrow denotes the location of ABL. (B) Heatmaps showing the phosphorylation of ABL substrates in men (top) and women (bottom) HCC tumor with Imatinib, Rebastinib, or Olverembatinib compared to vehicle. (C) The kinase activity of ABL and ARG/ABL2. The PerMed Score denote the percent change in kinase activity in the HCC tumor with ABL inhibitors compared to vehicle. The blue line denotes the average signal intensity of each substrate compared to the vehicle.

### Personalized Medicine Score Analysis

To determine how other kinase activities are affected by the three ABL antagonist treatments, we also quantified PerMed scores for other PTK and STK kinases. **Figures 7A and 7B** illustrate that adding ABL antagonists to the PamChip runs also changed other pathways, with some decreasing and some increasing. The inhibition of ABL kinase activity in these samples significantly increased ERK1 and AKT2 kinase activity, indicating that these pathways still actively signal and that changes are detectable in pathways by adding ABL inhibitors within the PamStation PamChip kinome runs. The strength of this analysis and the PerMed Score resides in their ability to predict whole kinome outcomes that lead to side effects in certain patient populations, and the sensitivity of the compound is measurable. Changes in some pathways were down in women’s HCC samples but elevated in men’s HCC samples, which could indicate hallmarks of drug responsiveness that may lead to sensitivity or resistance. Using this technology to calculate kinase activities of tens to hundreds of pathways offers a massive advantage in better profiling potential drug side effects and patient outcomes.

**Figure 7.**
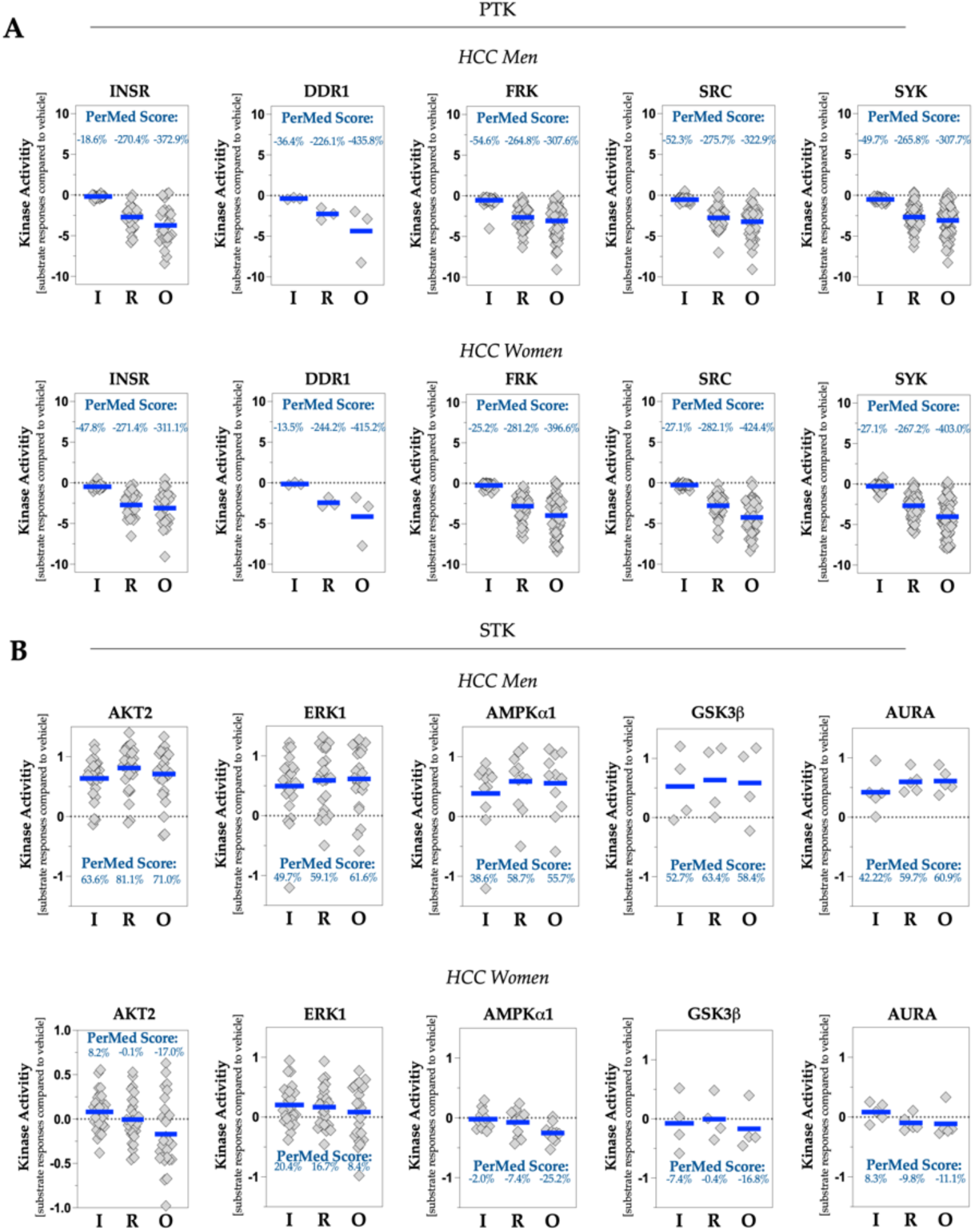
Off-target effects of ABL inhibitors using PerMed score analysis. (A) The kinase activity of PTK kinases INSR, DDR1, FRK, SRC, and SYK. The PerMed Score denote the percent change in kinase activity in the HCC tumor with ABL inhibitors compared to vehicle. The blue line denotes the average signal intensity of each substrate compared to the vehicle. (B) The kinase activity of STK kinases AKT2, ERK1, AMPKa1, GSK3b, and AURA. The PerMed Score denote the percent change in kinase activity in the HCC tumor with ABL inhibitors compared to vehicle. The blue line denotes the average signal intensity of each substrate compared to the vehicle.

These methods work for group analysis to find overall responses to a compound for drug discovery or individually for personalized medicine. Heatmap analysis of individual reactions to PTK and STK substrate phosphorylation indicates the efficacy of the compounds per patient sample (**Figure 8**).

**Figure 8.**
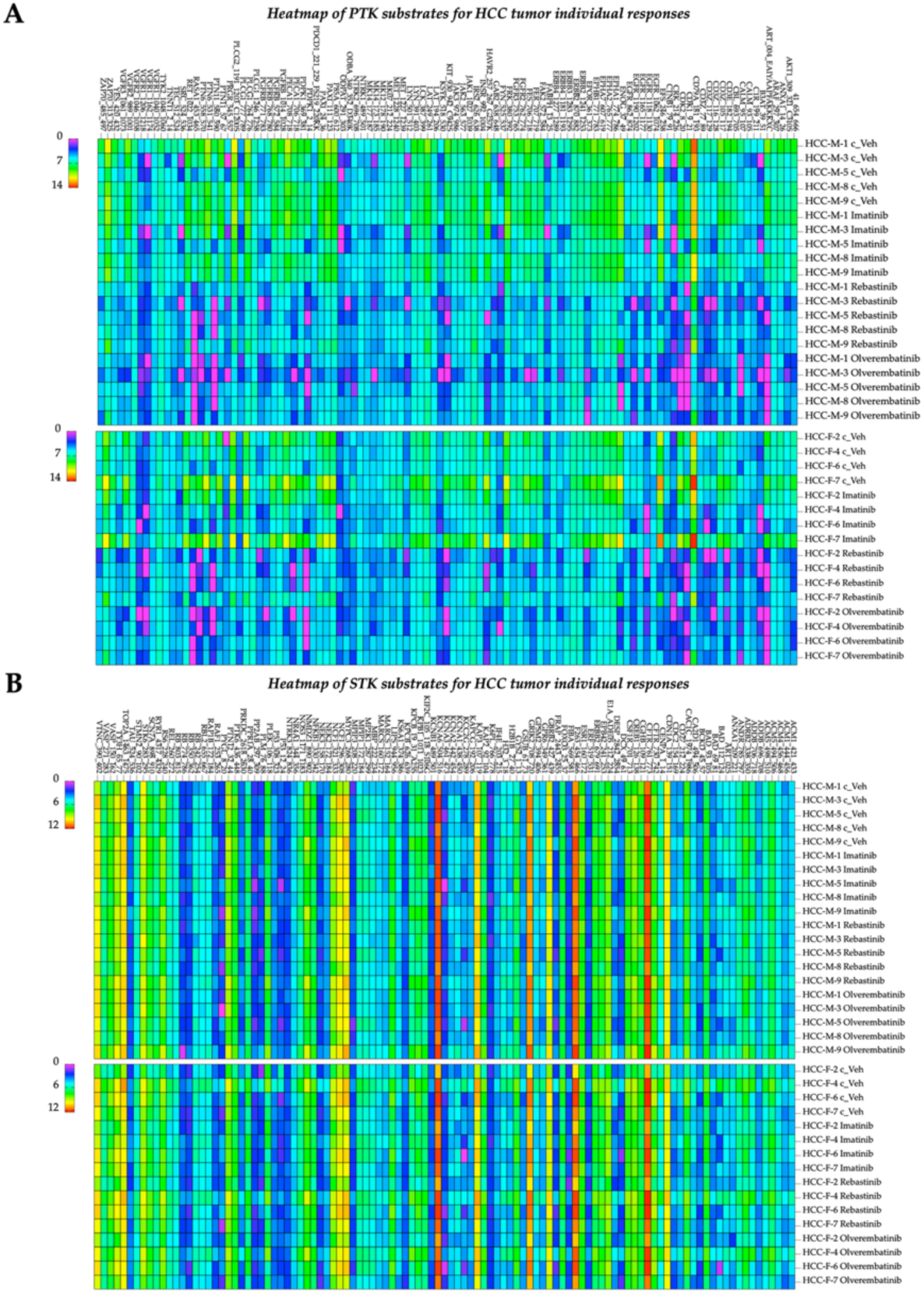
Effects of ABL antagonists on individual HCC samples. (A) Heatmaps showing the PTK substrates for HCC tumor responses from individual patients. (B) Heatmaps showing the STK substrates for HCC tumor responses from individual patients.

### Validation of Drug Actions on Kinases

To validate our findings with the HCC samples, we employed the human HCC HepG2 hepatocyte cell line, which was originally isolated from a liver biopsy of a Caucasian teenage male diagnosed with a well-differentiated tumor categorized as hepatoblastoma^24^. We extracted proteins from the HepG2 cells and introduced the same three ABL inhibitors to ascertain whether we could replicate our findings with human HCC samples. The PTK PamChip phosphorylation exhibited an almost identical pattern of responsiveness to the ABL antagonist when comparing the HepG2 cell proteins with HCC samples from both sexes (**Figure 9A**).

**Figure 9.**
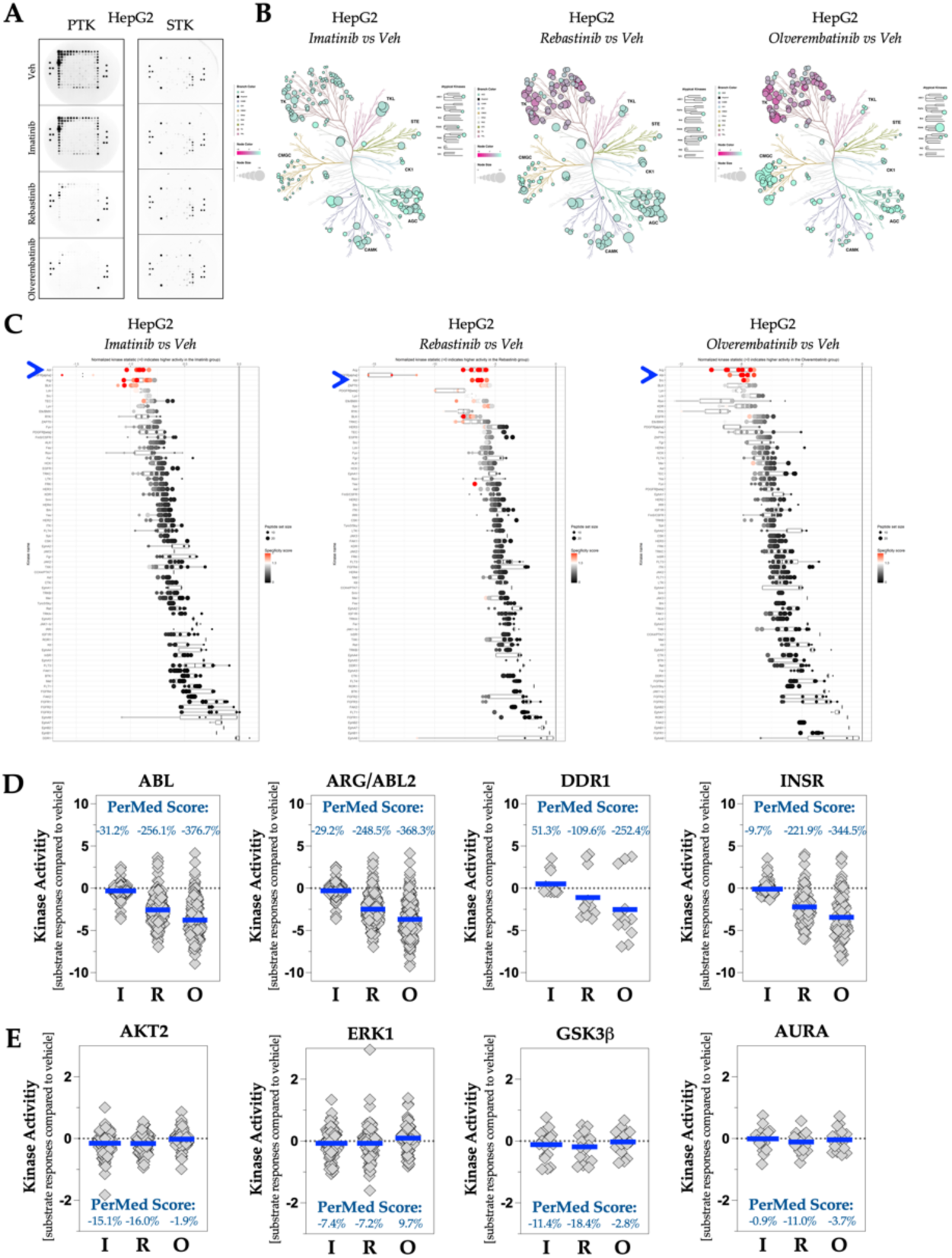
Validation of ABL antagonist actions on kinases. (A) Images of phosphorylated PTK (left) and STK (right) PamChip during the final cycle. (B) Phyla trees displaying the alterations of PTK and STK kinase activity in the HepG2 with Imatinib (left), Rebastinib (middle), or Olverembatinib (right) compared to vehicle. (C) Waterfall plot of the PTK Upstream Kinase Analysis (UKA) in HepG2 comparing Imatinib (left), Rebastinib (middle), and Olverembatinib (right) to vehicle. Blue arrow denotes the location of ABL. (D) The kinase activity of ABL, ARG/ABL2, DDR1, and INSR. The PerMed Score denote the percent change in kinase activity in the HepG2 with AB inhibitors compared to vehicle. The blue line denotes the average signal intensity of each substrate compared to the vehicle.

The visualization of HepG2 protein kinase activity responses to the ABL inhibitors, as represented in phylogenetic trees, also resembled the responsiveness observed in the HCC samples from men and women (**Figure 9B**). The BioNavigator upstream analysis identified ABL and ARG/ABL2 as the kinases targeted by the compounds, indicating inhibition of PTK kinases compared to vehicle treatments (**Figure 9C**). The PerMed score analysis for the PTK kinases reported -31.2% suppression for ABL and -29.2% for ARG/ABL2 when treated with imatinib (**Figure 9D**), which was more aligned with the responses of women’s HCC protein samples. Additionally, the DDR kinase activity exhibited an increase in response to imatinib; however, there was a decline with rebastinib and olverembatinib. The STK kinases displayed only modest variations in AKT2, ERK1, GSK3, and AURA activities (**Figure 9D**). Several STK kinases were suppressed following treatment with imatinib and rebastinib, whereas olverembatinib produced a divergent STK response, resulting in some kinases exhibiting increased activity while others demonstrated a decrease (**Supplemental Figure 4**).

By employing an additional four human hepatocyte cancer cell lines, we have corroborated these findings using male and female patients with HCC samples in conjunction with the analysis of HepG2 cell proteins. We observed that cells obtained from male and female subjects exhibited responses that were analogous to those observed in the kinase activities of the male and female HCC patient samples (**Supplemental Figures 5 & 6**). The ABL antagonists were tested using protein extracted from the four additional human hepatocyte cell lines, which include the human Hep3B2 HCC cell line derived from a young Black male, established concurrently with the human HepG2 HCC cells ^24^ (**Supplemental Figure 5**). Furthermore, we evaluated protein extracts from human HLE HCC cells sourced from a male patient 65-70 years old from Japan ^25^, Huh-7, which was ascertained from a Asian male patient 55-60 years old ^26^, and HepaRG cells derived from a tumor of a female patient from France, with HCC resulting from a hepatitis C infection ^27^ (**Supplemental Figures 5 & 6**). These results validate our findings from the HCC tumor protein lysates, which were processed individually for each patient sample and sex (**Supplemental Figures 2 & 3**).

Essentially, the ABL substrate specificity revealed the depth of the pathways detectable with specific proteins from each patient, which can be used to discern drug efficacy or disease pathological features. These technological advancements can be employed in software for an instrumented humanized ‘Clinical Trial in a PamChip’ or personal medicine (**Figure 10**), which can inform the patient about the best treatment options.

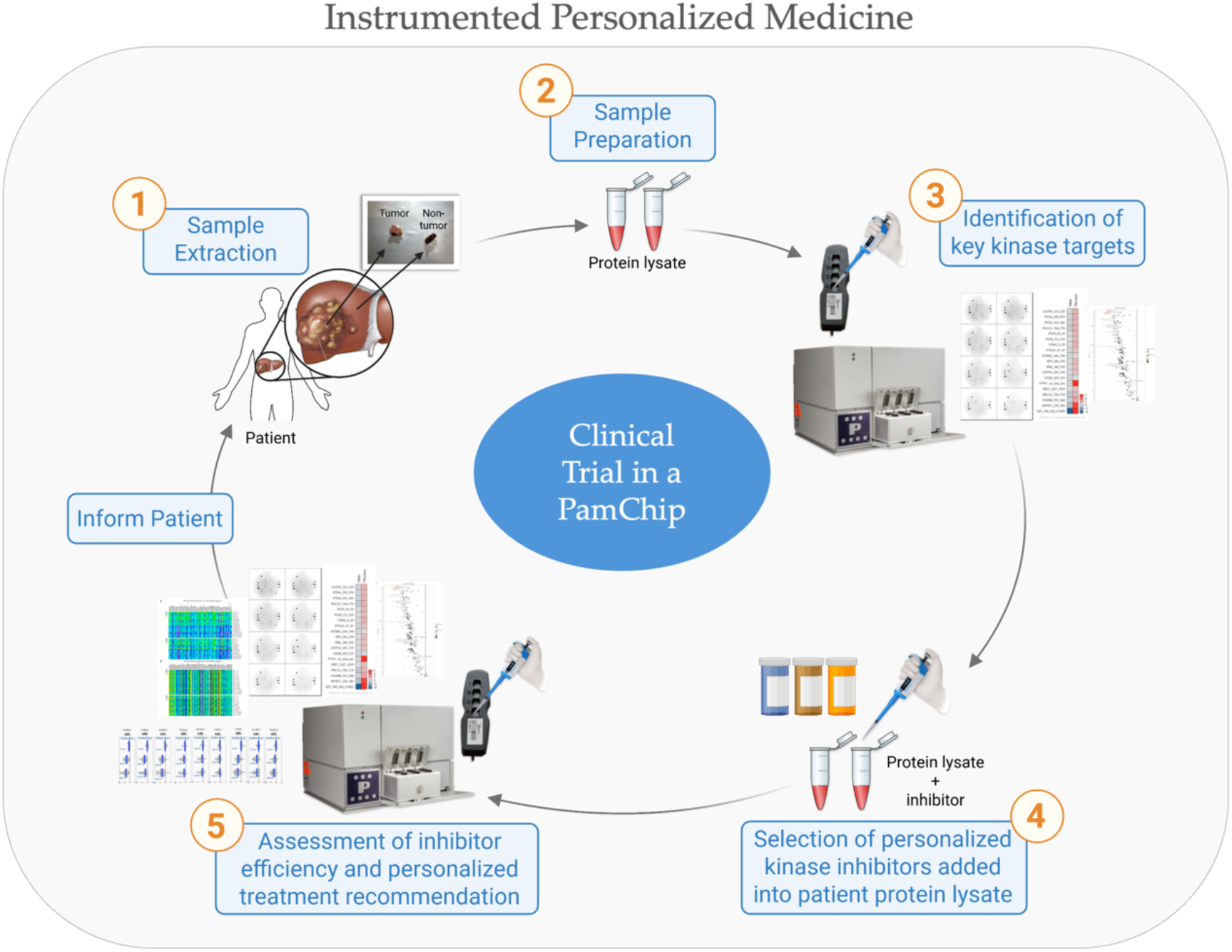

## DISCUSSION

In this work, we have made significant contributions to advancing kinome technology, which the PamStation holds considerable promise for the future of innovative personalized medicine and instrumented humanized clinical trials. This technology’s primary advantage is its ability to obtain signaling data from a patient’s biopsy or cells, thereby elucidating alterations in kinase activity. As demonstrated in our findings, this information could be employed to select the optimal medication for the individual. This process can apply multiple kinase inhibitors, which can be assessed utilizing the same lysate from the biopsied sample to identify the most effective drug for the patient. Implementing this approach can considerably reduce the time necessary to establish the appropriate treatment by precisely targeting the kinase pathways and identifying the most effective antagonist or agonist. Adopting PamGene technology in personalized medicine benefits nearly all patients and the healthcare system through its ability to measure hundreds of kinase activities for predictive tests, biomarkers, drug treatment responses, therapeutic development, and personalized medicine. The future of the PamGene PamStation technology is in its ability to predict signaling mechanisms in diseases and disorders and, most importantly, responses to treatments.

The observation that ABL was identified as the most active PTK through the Z-score waterfall plot is not unprecedented, as patients with HCC typically exhibit elevated ABL activity^21^. Several studies have also reported an increase in ABL expression; however, our analysis did not reveal a significantly increased expression but rather a modification in its kinase activity relative to target substrate proteins. Importantly, pathways that ABL inhibits, such as ERK1, AKT2, and AMPKα1, demonstrated heightened kinase activity in the presence of certain ABL antagonists. This indicates that these protein lysates maintain functional signaling capabilities despite being derived from tissue and cell lysates. The nine individual patient sample PamChip runs and the five human hepatocyte cancer cell lines demonstrated that off-target effects were quantifiable across hundreds of kinase signaling pathways. ABL antagonists have been documented to alter glucose sensitivity in some patients but not others. Gómez-Sámano *et al.* illustrated that individuals diagnosed with chronic myeloid leukemia or gastrointestinal stromal tumors, who also present with type 2 diabetes and are administered the ABL inhibitor imatinib, experienced a significant reduction in fasting plasma glucose and a decrease in glycosylated hemoglobin A1C levels ^28^. ERK1 and AKT2 are critical components of the insulin signaling cascade ^4^, and their increased signaling activity was evident with ABL inhibitors during the PamChip kinome assay, which is also valuable insight into off-target drug effects. Consequently, this data supports using the ‘Clinical Trial in a PamChip’ methodology to ascertain the efficacy of agonists and antagonists from a specific patient before treatment to detect off-target signaling accompanying side effects such as hypoglycemia. The effectiveness of each compound may be quantified, and the outcomes within the respective pathways can be assessed before patient administration.

Personalized medicine is critical in preventing, diagnosing, and treating diseases exhibiting diverse pathologies within the affected population. Omics techniques utilized in personalized medicine, such as DNA sequencing and its derivatives (RNA-seq, ChIP-seq, ATAC-seq, among others), have proven essential for comprehending the pathology of numerous diseases and metabolic states (metabolomics), which include various forms of cancer, MASLD, obesity, and cardiovascular diseases, and apply to nearly all disorders and diseases. Notwithstanding the advancements in RNA-seq and analogous technologies that report transcript abundance, these methods are limited in providing insights into functional pathways related to disease or drug treatment. Protein translation and activity are moderated by transcription factor activity ^29^, posttranslational modifications, and other regulatory mechanisms that influence protein stability where mRNA expression could be different from protein levels ^30^. For these reasons, establishing causal relationships by solely employing transcript abundance data derived from RNA-seq proves challenging and impractical in determining drug selectivity from patient samples. Our multi-omic kinome technique, designed to elucidate the complex mechanisms of disease and drug action, holds significant promise for research applications and has the potential to revolutionize clinical practices.

Omics data analysis constitutes the foundation of real-time kinase activity profiling with the PamGene. Given the nature of the measurement, a direct identification of which kinase contributes to the phosphorylation pattern of the 340 substrates is not readily accessible. Rather, it is derived through a pathway deconvolution process that generally aggregates all known peptide motifs that a kinase can phosphorylate. Subsequently, the data is compared to the phosphorylation pattern on the PamChip to ascertain the kinases most likely responsible for those phosphorylation events. The design of the PamGene PamChip kinome assays facilitates comprehensive omics data analysis, allowing users to assess protein functionality in diseases and drug therapies, particularly when multiple signaling pathways are disrupted.

The PamGene kinase activity profiling represents an innovative tool that has already demonstrated utility in research concerning cancers ^31–33^, liver fibrosis and cirrhosis ^10,18,19^, and metabolic diseases ^10,12,20,34–36^. In a prior investigation, we performed bioinformatic analyses of pathways in liver fibrosis, where we deconvoluted kinase signaling pathways using PamGene kinome and RNA-sequencing technologies. In separate publications using both technologies, we demonstrated that FOXS1 and INSR modulate fibrosis by regulating the levels of hepatic fibrotic signaling. ^18,19^ We utilized RNA-seq and the PamGene kinome analysis to elucidate the pathways they govern, and there were similarities among the findings in the two datasets. Others have profiled the kinome of peripheral blood mononuclear cells (PBMCs) derived from patients with melanoma or non-small cell lung cancer treated with two distinct immune checkpoint neutralizers^33^. The researchers successfully established a predictive model utilizing the kinome data to classify the treatment groups with an accuracy of up to 100% within the melanoma patient population ^33^. Additionally, other studies have assessed normal and malignant tissues from renal cell carcinoma patients and their responses to four different PTK inhibitors. They identified a set of 36 kinases that exhibited significantly differing activities between normal and cancerous tissues. Following *ex vivo* treatment of the samples, it was observed that tivozanib and cabozantinib exhibited a more pronounced inhibitory effect on the kinase pathways in cancerous tissues than sunitinib and pazopanib ^31^. Our study was not *ex vivo*, as the drugs were added directly to the reagent mixtures and adjusted for running the PamChip. Our study created a method many others can use to better understand and treat virtually any disease.

In this study, numerous members of the TK family exhibited hyperkinase activity in HCC of both sexes, as indicated by the PamGene technology. Our *Z*-score analysis in this study showed that ABL displayed the highest kinase activity in HCC in both sexes despite the unchanged mRNA levels. Most interesting and promising for personalized medicine is that the antagonism of ABL in the HCC samples not only led to the reversal of intended signaling pathways but also identified changes in other pathways that can be utilized as hallmarks of off-target signaling that led to drug side effects. In rare cases, it can identify a patient who doesn’t conventionally respond to the drug. This can be seen in Supplemental Figure 2B, in which patient HCC-F-7 displayed increased ABL activity with imatinib. This surprising data point could enormously impact the patient’s treatment. If treated with imatinib, the patient’s condition may worsen. This technology allows the unique ability to foresee patient response to treatment and, therefore, can facilitate treatment plans, improving the patient’s chances of success.

Here, we present a kinomic atlas for human HCC tumors, validating our findings in human HCC samples and cells. This endeavor may significantly enhance the scientific understanding of the mechanisms underlying HCC. We have expanded the capabilities of the PamGene PamStation technology to incorporate drug compounds, such as ABL antagonists, and this method can also be used to test other compounds in different diseases and tissues. When we combined the HCC samples with the ABL inhibitors and re-ran the same sample lysates, we detected significantly less ABL kinase activity with the antagonists present. Even though there was no intact cell signaling system, the samples consisted of lysates with residual kinase activity remaining. The bioinformatics analyses demonstrated the effectiveness and sensitivity of these compounds, revealing alterations in numerous pathways despite the absence of intact cells. Observing an overactive ABL kinase in HCC was not novel; however, the concurrent utilization of ABL antagonists while examining the PamChip kinome within these samples has not been previously conducted.

In conclusion, the PamStation kinome technology exhibits considerable clinical usability; nonetheless, its implementation in clinical environments is restricted due to cost, usability, and data analysis challenges. The PamGene PamStation PamChip technology presents numerous practical applications for understanding diseases, drug actions, and personalized medicine. Our methodology, centered on instrumented personalized medicine, enhances the capacity to classify patient subtypes based on their kinase signaling pathways, which could improve both responsiveness and sensitivity to treatment. Our advancement of the PamGene kinase technology may serve as a diagnostic tool for identifying the causes and mechanisms of diseases. Artificial intelligence (AI) software could be used to identify patterns in the changes of all kinases using the PerMed Score. This software can advance disease understanding, drug action, side effects, and many other benefits that can improve clinical outcomes. Ultimately, this could lead to more effective therapeutics tailored to an individual’s protein responsiveness, enhancing both the patient’s experience and the overall success of treatment

## METHODS

### Human Subjects

The Institutional Review Board (IRB) at the University of Kentucky waived ethical approval for this work as the human liver samples were deidentified, and the authors were not given access to any patient identifiers. The sample names were randomly assigned to ensure the results could not be linked to a patient. The University of Kentucky Institutional Review Board (IRB) deemed this project did not require full IRB review because it fulfills the “United States Department of Health & Human Services 2008 Coded Private Information or Specimen Use in Research Guidance” requirements for this decision. All other methods and procedures are described in more detail in the STAR★METHODS

## Resource availability

### Lead Contact

Requests for resources and further information should be directed to the lead contact, Dr. Terry Hinds.

### Materials Availability

This study did not generate any new unique reagents.

### Data and Code Availability

The kinase reports and raw files are available on *TBD* at the publication date. This study did not generate any unique code.

## Data Availability

All data produced in the present work are contained in the manuscript

## Acknowledgments

This work was supported by grants from the National Institutes of Health (NIH) R01DK121797 (T.D.H.J.), R01DA058933 (T.D.H.J.), R01HL174521 (T.D.H.J.), F31HL170972 (Z.A.K.), F31HL175979 (E.A.B.), and American Heart Association (AHA) grant 25PRE1374495 (G.J.M.). The HCC and normal adjacent tissue were received from and supported by the Biospecimen Procurement & Translational Pathology Shared Resource Facility of the University of Kentucky Markey Cancer Center (P30CA177558). The contents are solely the responsibility of the authors and do not necessarily represent the official views of the NIH or AHA. The authors thank Matthew Hazzard and Thomas Dolan of the University of Kentucky College of Medicine for generating Figure 10. The authors thank Dr. Robert Helsley for the HLE cells, Dr. Sadeesh Ramakrishnan at the University of Pittsburgh for the Huh-7 cells, and Dr. Samir Softic while they were at the University of Kentucky (currently at Pikeville Medical Center) for HepG2 cells.

## Author contributions

Conceptualization, Z.A.K., E.A.B., and T.D.H.J.; Data Curation, Z.A.K. and E.A.B.; Formal Analysis, Z.A.K., E.A.B., G.J.M., S.N.P., and T.D.H.J.; Funding Acquisition, Z.A.K., E.A.B., G.J.M., and T.D.H.J.; Investigation, Z.A.K., E.A.B., G.J.M., W.H.L., and S.N.P.; Methodology, Z.A.K. and T.D.H.J.; Project Administration, Z.A.K. and T.D.H.J.; Resources, T.D.H.J.; Supervision; T.D.H.J.; Validation, Z.A.K., E.A.B., G.J.M., W.H.L., and S.N.P.; Visualization, Z.A.K., E.A.B., G.J.M., W.H.L., and T.D.H.J.; Writing - original draft, Z.A.K. and T.D.H.J.; Writing – review & editing, Z.A.K., E.A.B., G.J.M., W.H.L., S.N.P., and T.D.H.J.

## SUPPLEMENTAL TABLES

**Supplemental Table 1.**
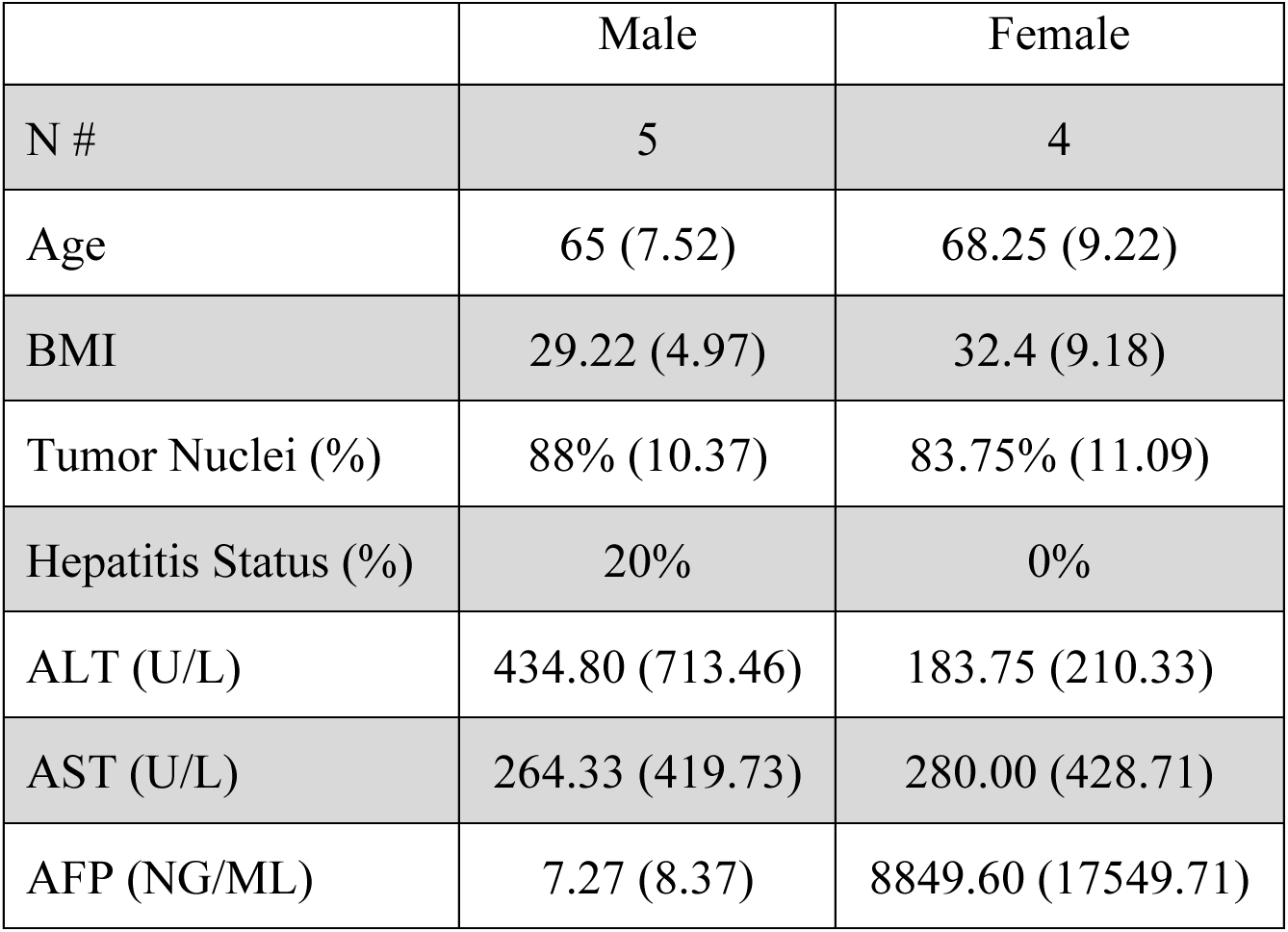
Liver Biopsy Patient Clinical Characteristics (expressed as: average (standard deviation).

|  | Male | Female |
| --- | --- | --- |
| N # | 5 | 4 |
| Age | 65 (7.52) | 68.25 (9.22) |
| BMI | 29.22 (4.97) | 32.4 (9.18) |
| Tumor Nuclei (%) | 88% (10.37) | 83.75% (11.09) |
| Hepatitis Status (%) | 20% | 0% |
| ALT (U/L) | 434.80 (713.46) | 183.75 (210.33) |
| AST (U/L) | 264.33 (419.73) | 280.00 (428.71) |
| AFP (NG/ML) | 7.27 (8.37) | 8849.60 (17549.71) |

**Supplemental Table 2A.** Phospho-Tyrosine Kinase (PTK) - Male HCC vs Male Adjacent Normal Upstream Kinase Analysis (UKA) - Kinase Families.

| Kinase | Observed | SamplingAvg | SD | Z |
| --- | --- | --- | --- | --- |
| ABL | 43 | 36.582 | 3.203 | 2.004 |
| PDGFR | 84 | 77.930 | 3.086 | 1.967 |
| SRC | 97 | 92.185 | 2.634 | 1.828 |
| SYK | 79 | 73.218 | 3.396 | 1.703 |
| JAK | 14 | 18.267 | 2.530 | -1.686 |
| FAK | 37 | 32.522 | 3.088 | 1.450 |
| TEC | 84 | 79.702 | 3.093 | 1.389 |
| ALK | 25 | 28.969 | 2.988 | -1.328 |
| DDR | 24 | 20.661 | 2.594 | 1.287 |
| RET | 5 | 7.064 | 1.636 | -1.261 |
| INSR | 22 | 18.952 | 2.513 | 1.213 |
| EGFR | 72 | 76.162 | 3.454 | -1.205 |
| CSK | 44 | 40.698 | 3.234 | 1.021 |
| AXL | 37 | 34.190 | 3.053 | 0.921 |
| ACK | 1 | 0.586 | 0.493 | 0.841 |
| SEV | 1 | 0.588 | 0.492 | 0.837 |
| EPH | 20 | 21.850 | 2.766 | -0.669 |
| VEGFR | 17 | 18.375 | 2.516 | -0.546 |
| FER | 6 | 5.320 | 1.450 | 0.469 |
| MET | 19 | 20.160 | 2.599 | -0.446 |

**Supplemental Table 2B.** Phospho-Tyrosine Kinase (PTK) - Female HCC vs Female Adjacent Normal Upstream Kinase Analysis (UKA) - Kinase Families.

| **Kinase** | **Observed** | **SamplingAvg** | **SD** | **Z** |
| --- | --- | --- | --- | --- |
| INSR | 19 | 12.743 | 2.549 | 2.455 |
| **ABL** | 31 | 24.720 | 3.128 | 2.008 |
| SYK | 56 | 49.556 | 3.325 | 1.938 |
| SRC | 59 | 62.264 | 2.705 | -1.206 |
| FER | 2 | 3.595 | 1.449 | -1.100 |
| FRK | 7 | 5.150 | 1.719 | 1.076 |
| AXL | 20 | 23.146 | 3.052 | -1.031 |
| TRK | 6 | 7.987 | 2.073 | -0.959 |
| ACK | 0 | 0.401 | 0.490 | -0.818 |
| SEV | 0 | 0.390 | 0.488 | -0.800 |
| DDR | 12 | 14.050 | 2.670 | -0.768 |
| PDGFR | 55 | 52.594 | 3.161 | 0.761 |
| EGFR | 54 | 51.612 | 3.361 | 0.711 |
| VEGFR | 14 | 12.456 | 2.541 | 0.608 |
| FAK | 20 | 21.837 | 3.110 | -0.591 |
| MET | 12 | 13.518 | 2.577 | -0.589 |
| JAK | 11 | 12.363 | 2.518 | -0.541 |
| RET | 4 | 4.826 | 1.658 | -0.498 |
| EPH | 16 | 14.780 | 2.774 | 0.440 |
| TEC | 53 | 53.846 | 3.194 | -0.265 |
| FGFR | 21 | 20.270 | 2.944 | 0.248 |
| RYK | 1 | 0.831 | 0.695 | 0.243 |
| ALK | 19 | 19.549 | 2.955 | -0.186 |
| CSK | 27 | 27.568 | 3.312 | -0.171 |

**Supplemental Table 3A.** Serine-Threonine Kinase (STK) - Male HCC vs Male Adjacent Normal Upstream Kinase Analysis (UKA) - Kinase Families.

| Kinase | Observed | SamplingAvg | SD | Z |
| --- | --- | --- | --- | --- |
| HCK | 34 | 26.059 | 2.896 | 2.742 |
| Yes | 31 | 23.542 | 2.745 | 2.717 |
| HER2 | 28 | 36.551 | 3.211 | -2.663 |
| ALK | 40 | 47.320 | 3.273 | -2.237 |
| FRK | 51 | 43.596 | 3.337 | 2.219 |
| Tyk2 | 1 | 3.558 | 1.164 | -2.198 |
| Lck | 31 | 24.721 | 2.862 | 2.194 |
| JAK1 | 1 | 3.562 | 1.173 | -2.184 |
| EphB1 | 1 | 3.538 | 1.174 | -2.162 |
| Lyn | 23 | 17.616 | 2.591 | 2.078 |
| FAK1 | 41 | 47.910 | 3.337 | -2.071 |
| BLK | 27 | 21.772 | 2.682 | 1.949 |
| HER4 | 29 | 34.742 | 3.041 | -1.888 |
| EphA7 | 1 | 2.990 | 1.076 | -1.850 |
| FGFR3 | 9 | 12.993 | 2.167 | -1.842 |
| Src | 40 | 34.205 | 3.194 | 1.814 |
| EphB2 | 1 | 2.973 | 1.114 | -1.770 |
| EphB3 | 1 | 2.934 | 1.093 | -1.770 |
| EGFR | 36 | 41.980 | 3.394 | -1.762 |
| JAK3 | 3 | 5.329 | 1.405 | -1.658 |
| Ros | 4 | 2.394 | 1.001 | 1.605 |
| KDR | 13 | 16.539 | 2.428 | -1.458 |
| FGFR2 | 11 | 14.190 | 2.257 | -1.413 |
| FLT4 | 10 | 12.985 | 2.149 | -1.389 |
| GSK3[beta] | 0 | 0.615 | 0.487 | -1.264 |

**Supplemental Table 3B.** Serine-Threonine Kinase (STK) - Female HCC vs Female Adjacent Normal Upstream Kinase Analysis (UKA) - Kinase Families.

| Kinase | Observed | SamplingAvg | SD | Z |
| --- | --- | --- | --- | --- |
| Mer | 23 | 31.062 | 3.343 | -2.412 |
| FLT3 | 7 | 12.314 | 2.440 | -2.178 |
| FAK1 | 25 | 32.414 | 3.443 | -2.153 |
| Tyk2 | 0 | 2.398 | 1.185 | -2.023 |
| JAK1 | 0 | 2.381 | 1.191 | -1.999 |
| Met | 18 | 24.009 | 3.187 | -1.885 |
| EphB4 | 0 | 1.998 | 1.077 | -1.855 |
| JAK3 | 1 | 3.586 | 1.415 | -1.827 |
| MAP2K7 | 2 | 0.758 | 0.690 | 1.801 |
| MEK1/MAP2K1 | 2 | 0.798 | 0.692 | 1.736 |
| SEK1/MAP2K4 | 2 | 0.792 | 0.701 | 1.724 |
| Axl | 32 | 37.358 | 3.497 | -1.532 |
| FAK2 | 17 | 21.512 | 3.079 | -1.465 |
| Fyn | 7 | 10.348 | 2.351 | -1.424 |
| Lmr1 | 0 | 1.202 | 0.861 | -1.397 |
| Kit | 9 | 12.413 | 2.469 | -1.382 |
| ASK/MAP3K5 | 1 | 0.372 | 0.483 | 1.299 |
| Src | 19 | 23.164 | 3.236 | -1.287 |
| CDK2 | 1 | 0.376 | 0.485 | 1.287 |
| Wee2 | 1 | 0.384 | 0.486 | 1.268 |
| CDC2/CDK1 | 1 | 0.387 | 0.487 | 1.258 |
| ERK2 | 1 | 0.388 | 0.487 | 1.257 |
| RAF1 | 1 | 0.388 | 0.487 | 1.256 |
| FGFR3 | 6 | 8.711 | 2.161 | -1.255 |
| MKK6/MAP2K6 | 1 | 0.390 | 0.488 | 1.250 |

**Supplemental Table 4.**
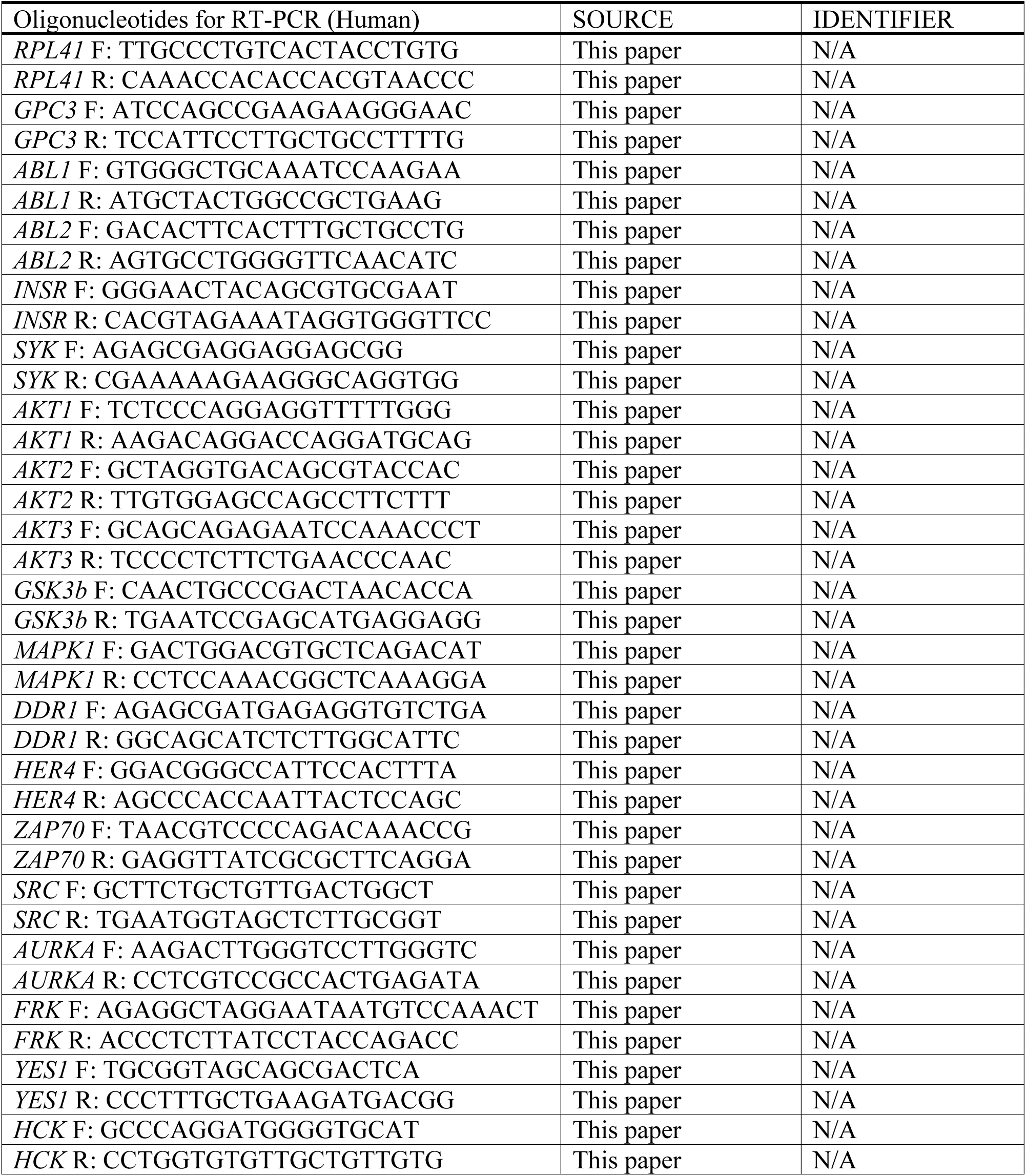
Human Real-Time PCR Primers.

## SUPPLEMENTAL FIGURES

**Figure S1.**
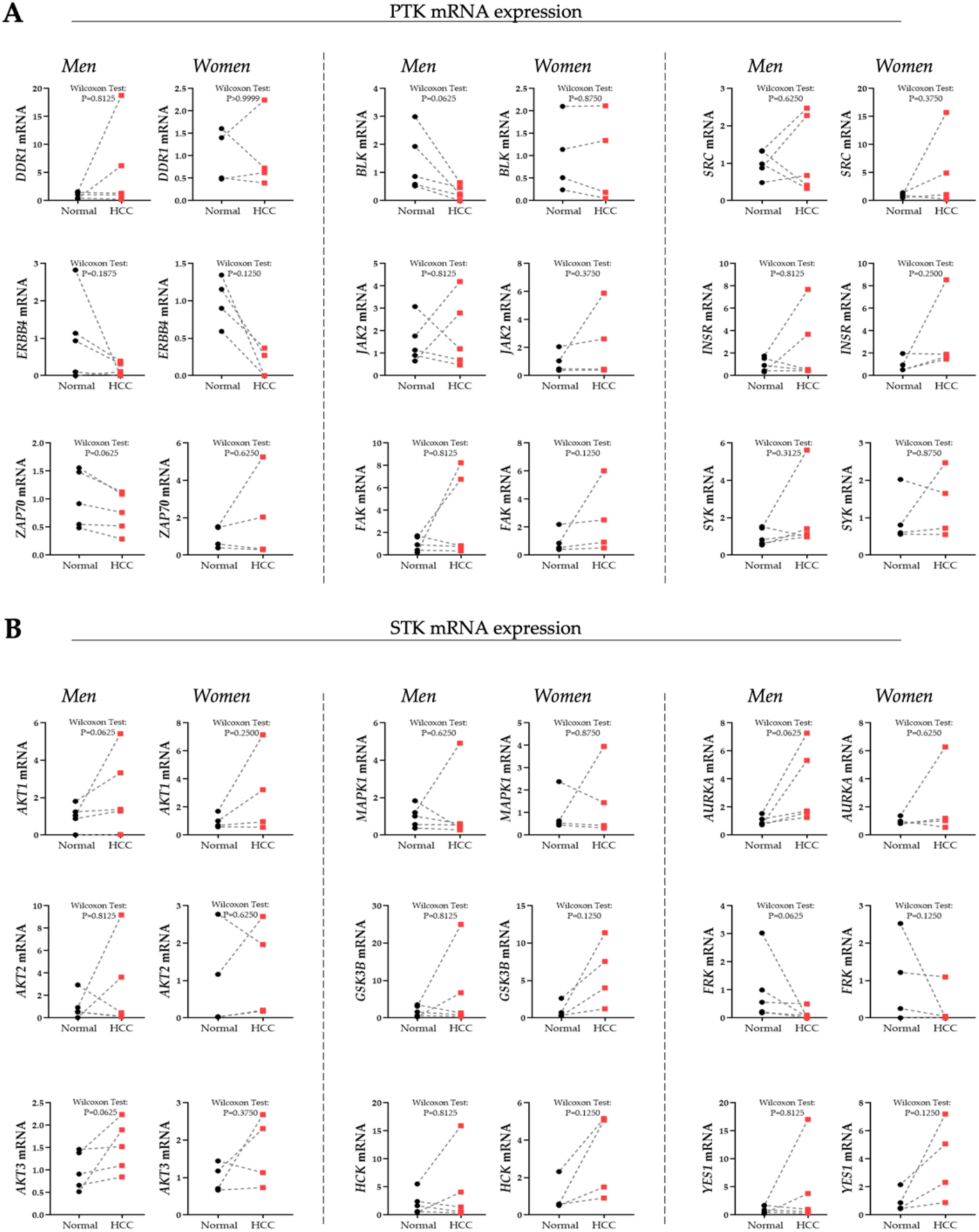
RT-PCR validation of altered kinases. RT-PCR validation of PTK (A) and STK (B) kinases uncovered in the kinome analysis in the men and women HCC tumor and normal adjacent tissue samples.

**Figure S2.**
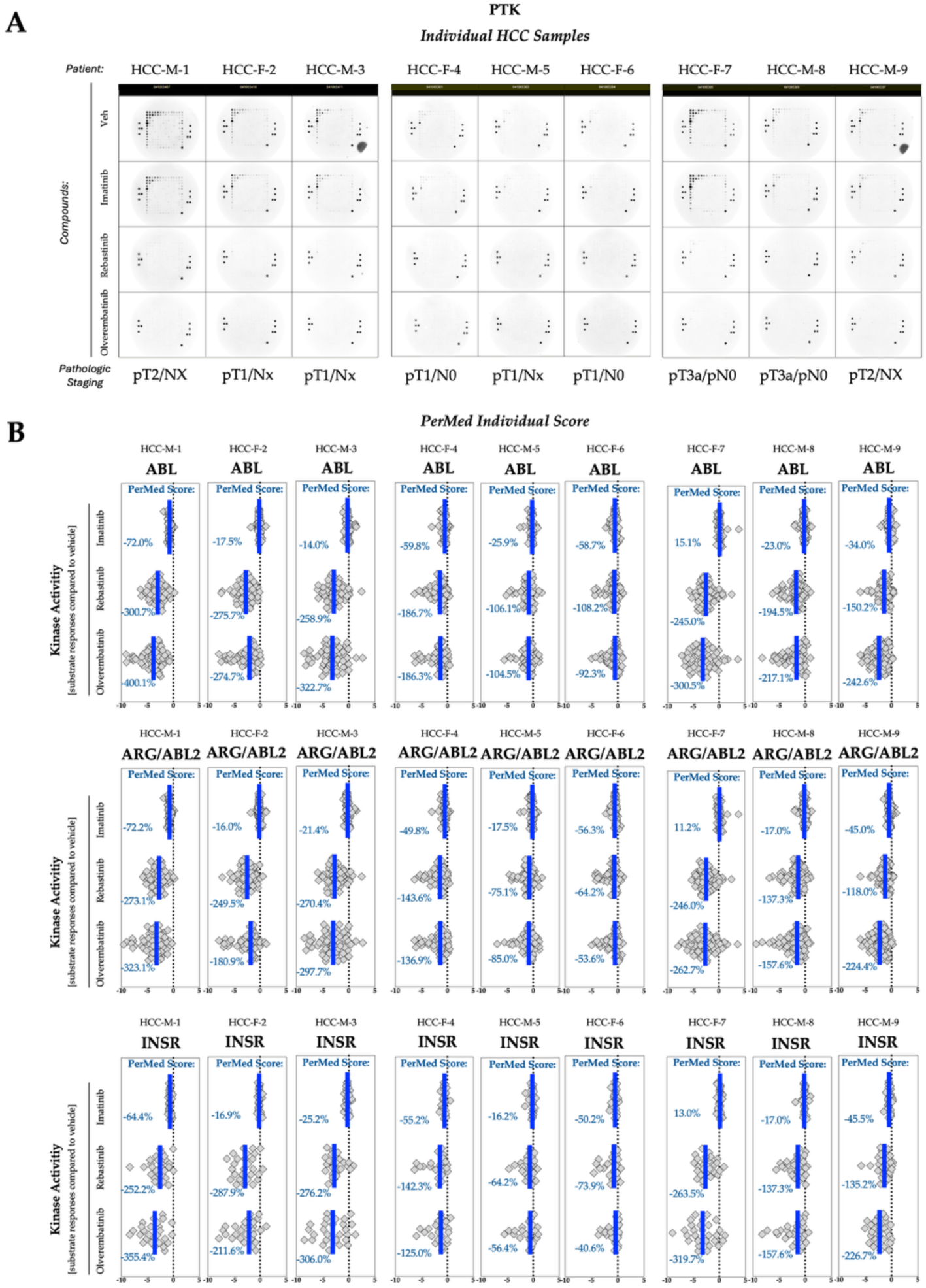
The efficacy of ABL inhibitors in nine HCC tumors. (A) Images of phosphorylated PTK PamChip during the final cycle. The tumor stage is denoted on the bottom. (B) The kinase activity of ABL, ARG/ABL2, and INSR. The PerMed Score denote the percent change in kinase activity in the HCC tumor with ABL inhibitors compared to vehicle. The blue line denotes the average signal intensity of each substrate compared to the vehicle.

**Figure S3.**
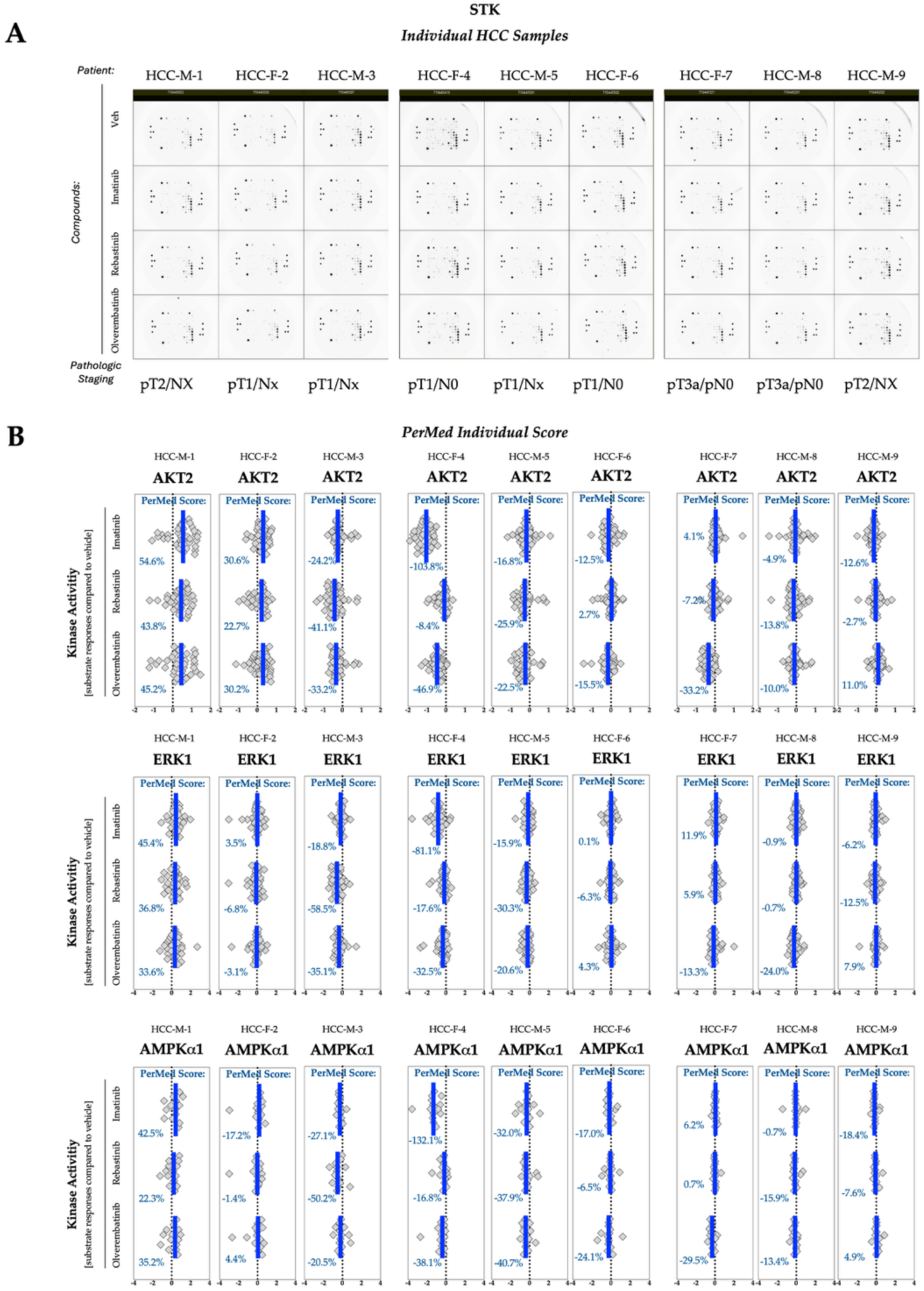
Off-target effects on STK kinases of ABL inhibitors in nine HCC tumors. (A) Images of phosphorylated STK PamChip during the final cycle. The tumor stage is denoted on the bottom. (B) The kinase activity of AKT2, ERK1, and AMPKa1. The PerMed Score denote the percent change in kinase activity in the HCC tumor with ABL inhibitors compared to vehicle. The blue line denotes the average signal intensity of each substrate compared to the vehicle.

**Figure S4.**
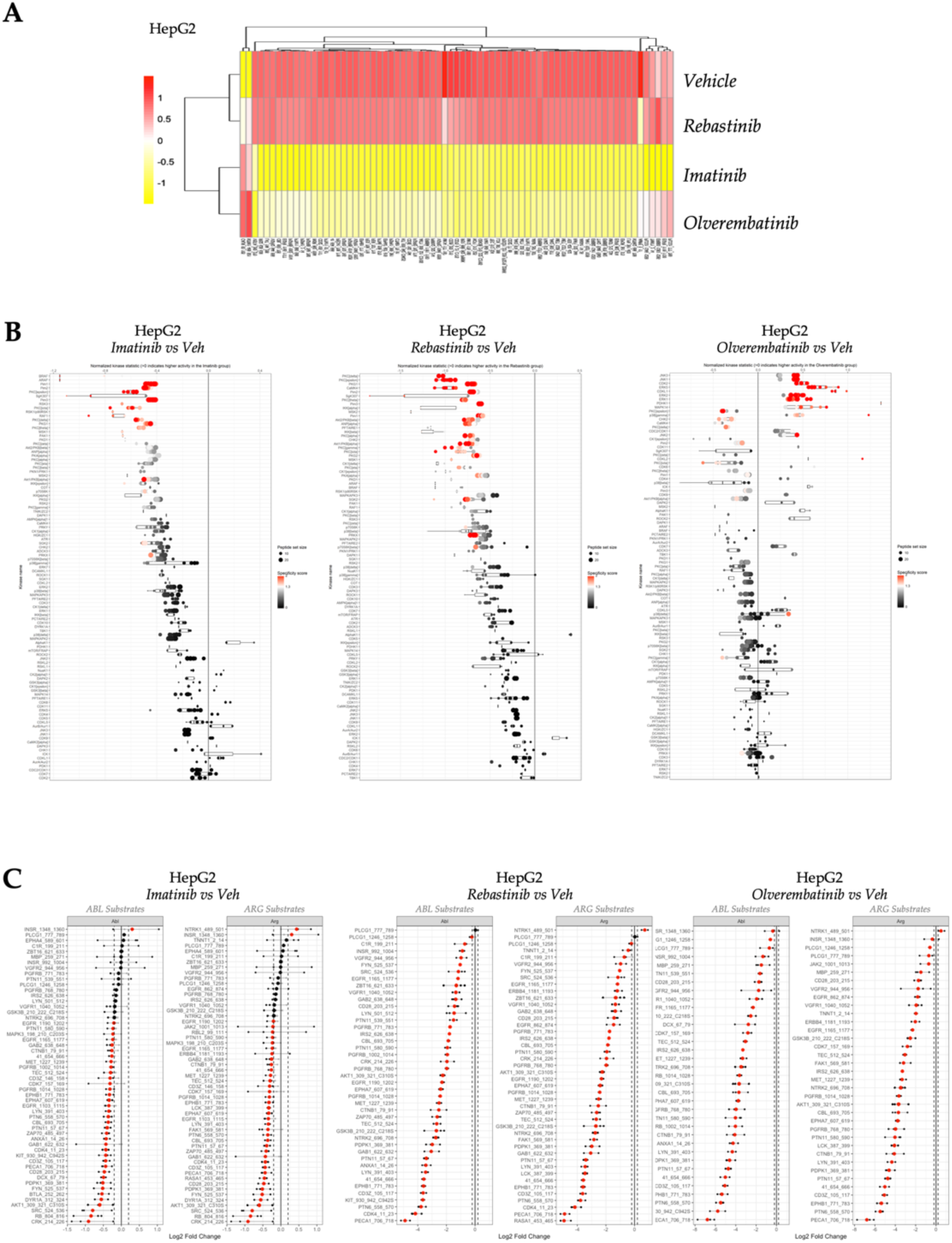
ABL inhibitors differentially suppress PTK and STK kinases in HepG2. (A) Heatmap showing the phosphorylation of 196 PTK substrates in HepG2 with vehicle, Imatinib, Rebastinib, or Olverembatinib. (B) Waterfall plot of the STK Upstream Kinase Analysis (UKA) in HepG2 comparing Imatinib (left), Rebastinib (middle), and Olverembatinib (right) to vehicle. (C) Waterfall plots of ABL and ARG substrates in HepG2 with Imatinib (left), Rebastinib (middle), and Olverembatinib (right) compared to vehicle.

**Figure S5.**
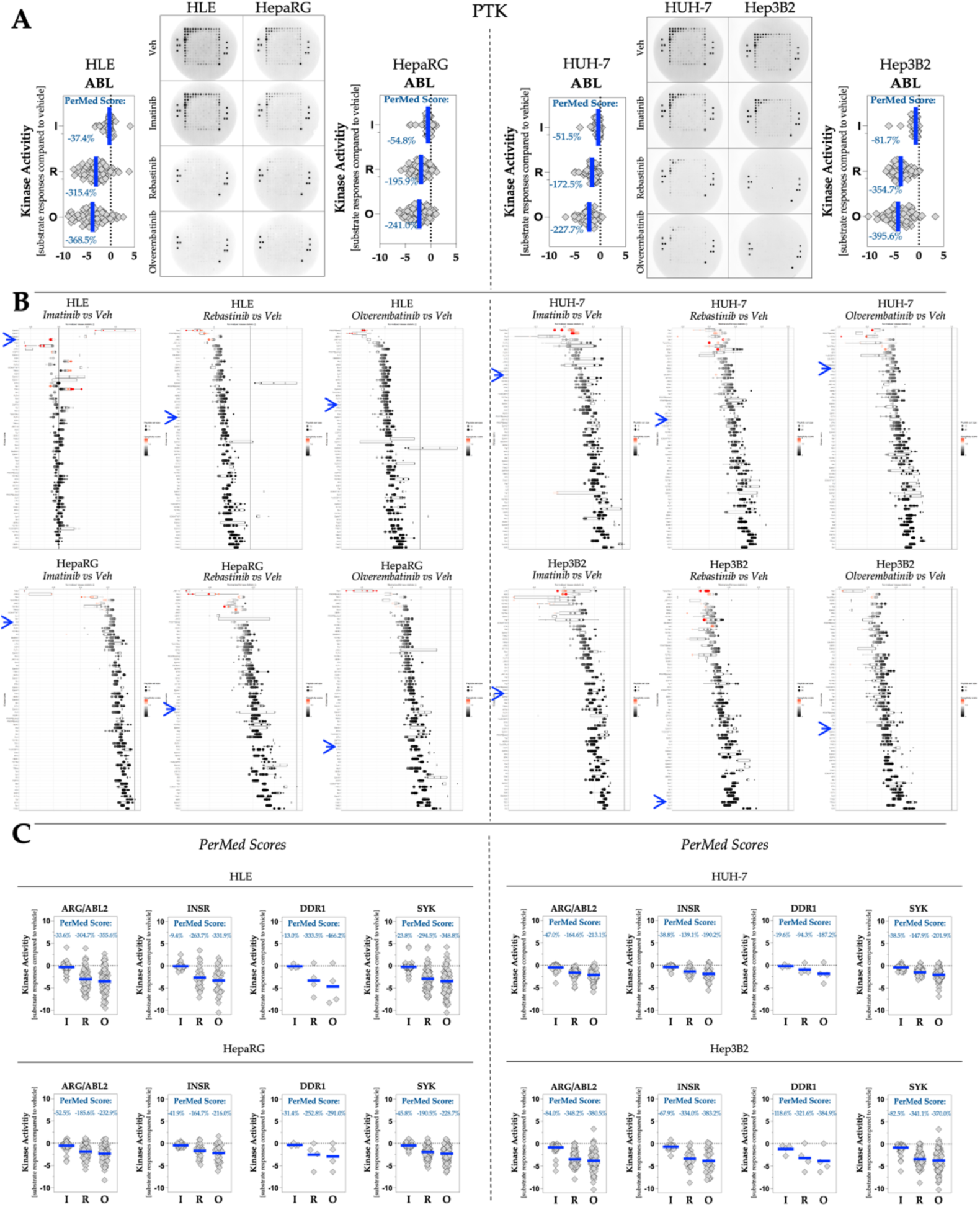
ABL inhibitors suppress PTK kinases in human hepatocyte cancer cell lines. (A) Images of phosphorylated PTK PamChip during the final cycle in HLE, HepaRG, Huh7, and Hep3B2 cells. The kinase activity of ABL in the four cell lines. The PerMed Score denote the percent change in kinase activity in the cell lines with ABL inhibitors compared to vehicle. The blue line denotes the average signal intensity of each substrate compared to the vehicle. (B) Waterfall plot of the PTK Upstream Kinase Analysis (UKA) in HLE, HepaRG, Huh7, and Hep3B2 cells comparing Imatinib (left), Rebastinib (middle), and Olverembatinib (right) to vehicle. Blue arrow denotes the location of ABL. (C) The kinase activity of ARG/ABL2, INSR, DDR1, and SYK. The PerMed Score denote the percent change in kinase activity in the cell lines with ABL inhibitors compared to vehicle. The blue line denotes the average signal intensity of each substrate compared to the vehicle.

**Figure S6.**
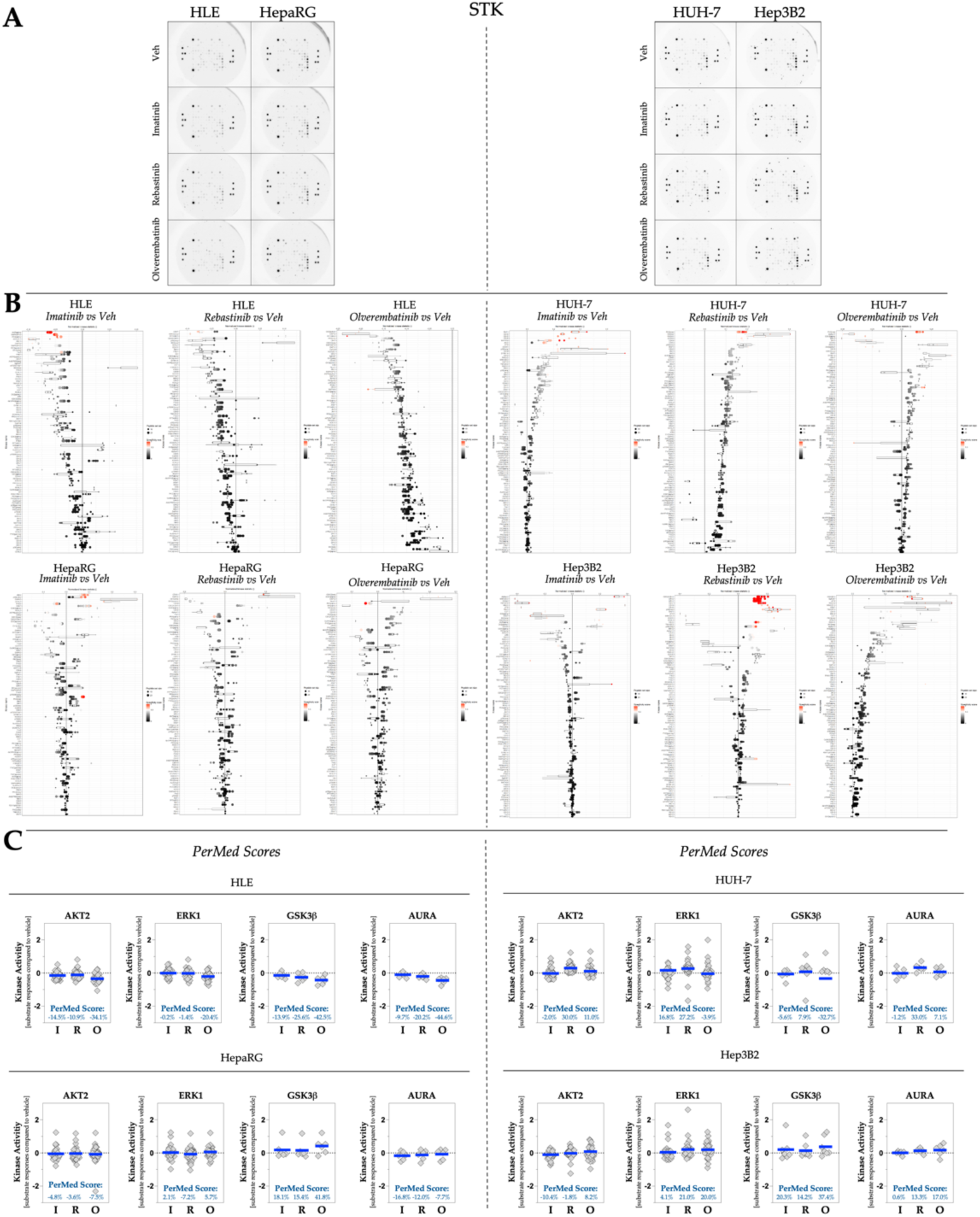
ABL inhibitors differentially suppress STK kinases in human hepatocyte cancer cell lines. (A) Images of phosphorylated STK PamChip during the final cycle in HLE, HepaRG, Huh7, and Hep3B2 cells. (B) Waterfall plot of the STK Upstream Kinase Analysis (UKA) in HLE, HepaRG, Huh7, and Hep3B2 cells comparing Imatinib (left), Rebastinib (middle), and Olverembatinib (right) to vehicle. (C) The kinase activity of AKT2, ERK1, GSK3b, and AURA. The PerMed Score denote the percent change in kinase activity in the cell lines with ABL inhibitors compared to vehicle. The blue line denotes the average signal intensity of each substrate compared to the vehicle.

## Appendix A: Individual HCC Samples with ABL Inhibitors

### PTK PamChip runs

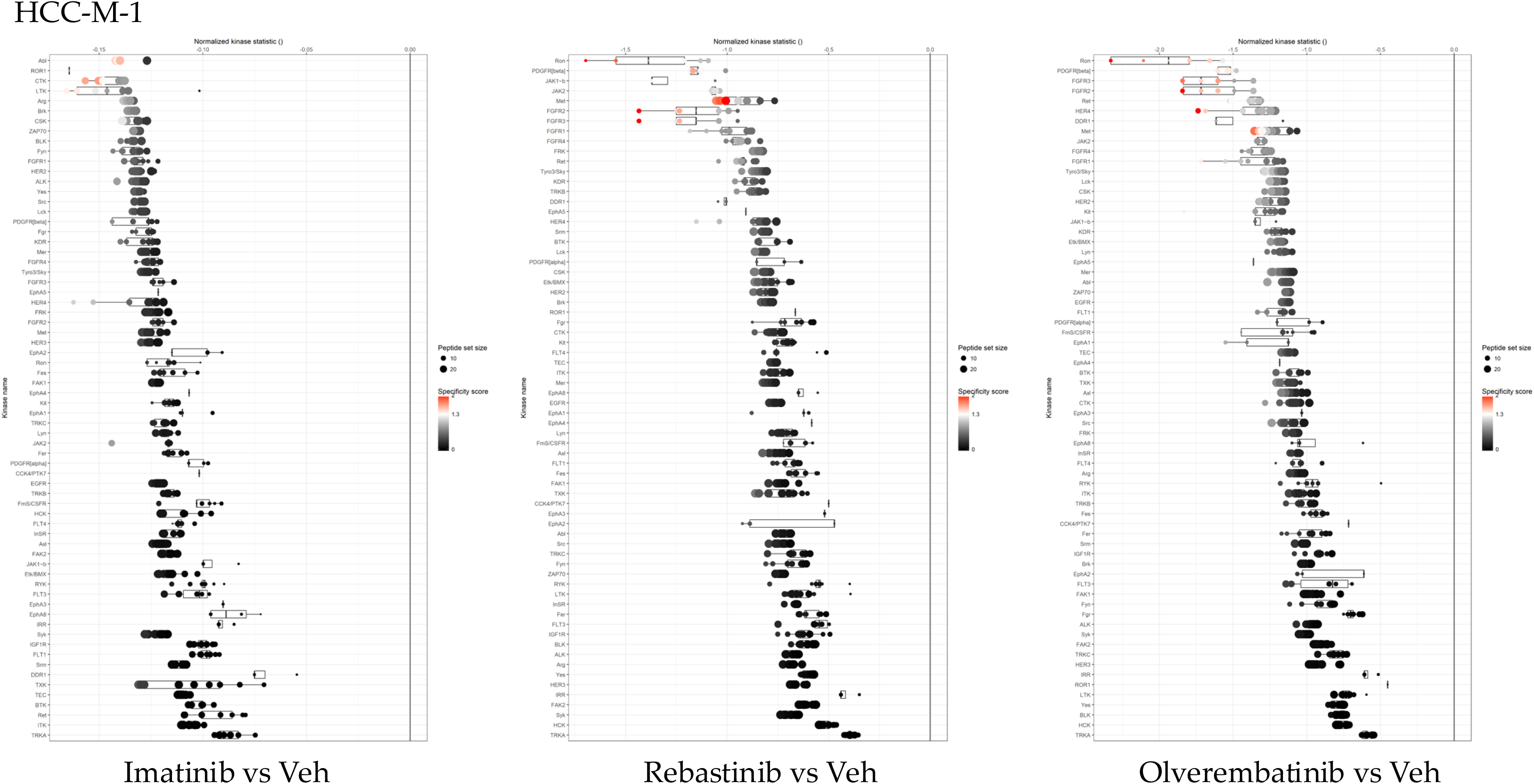
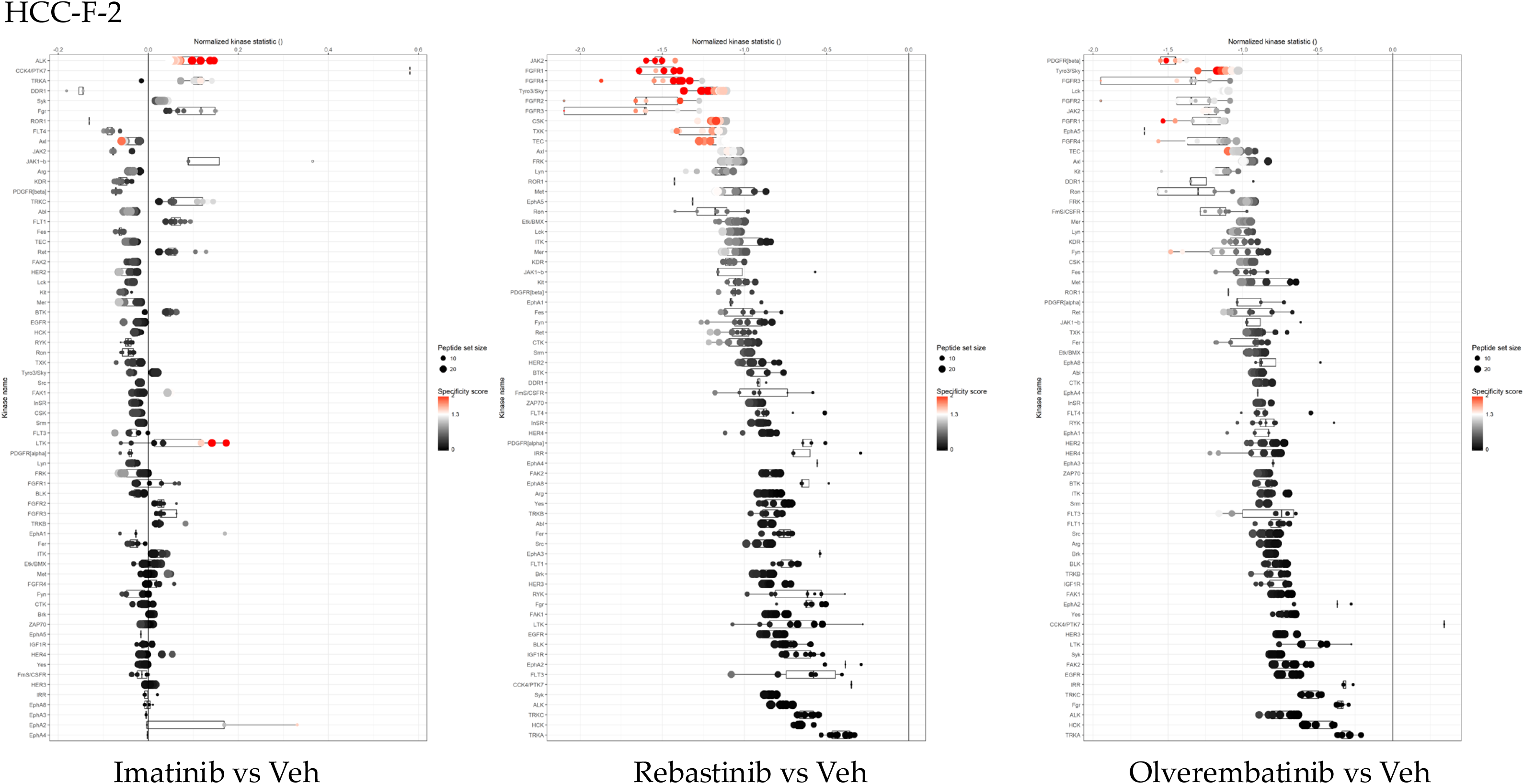
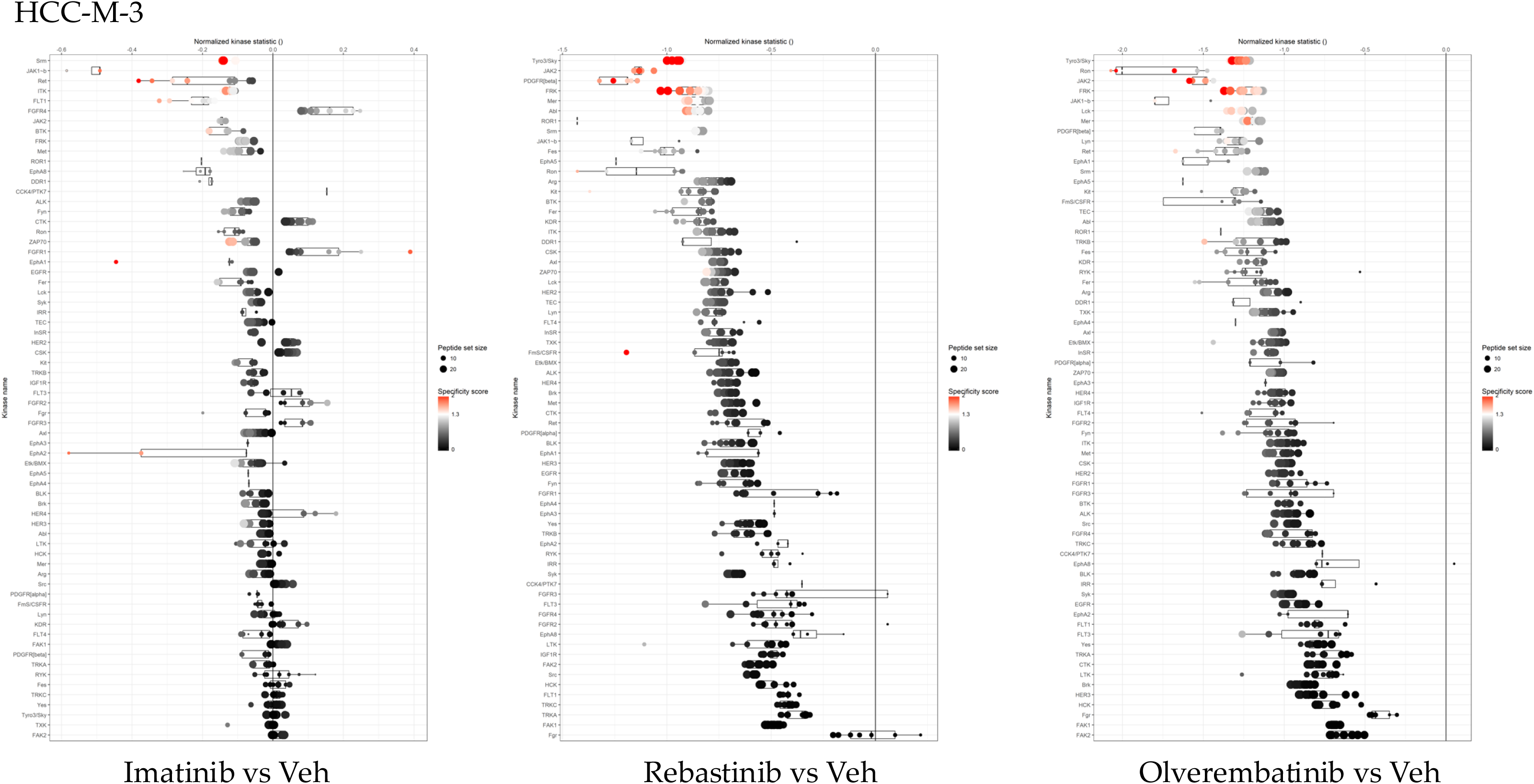
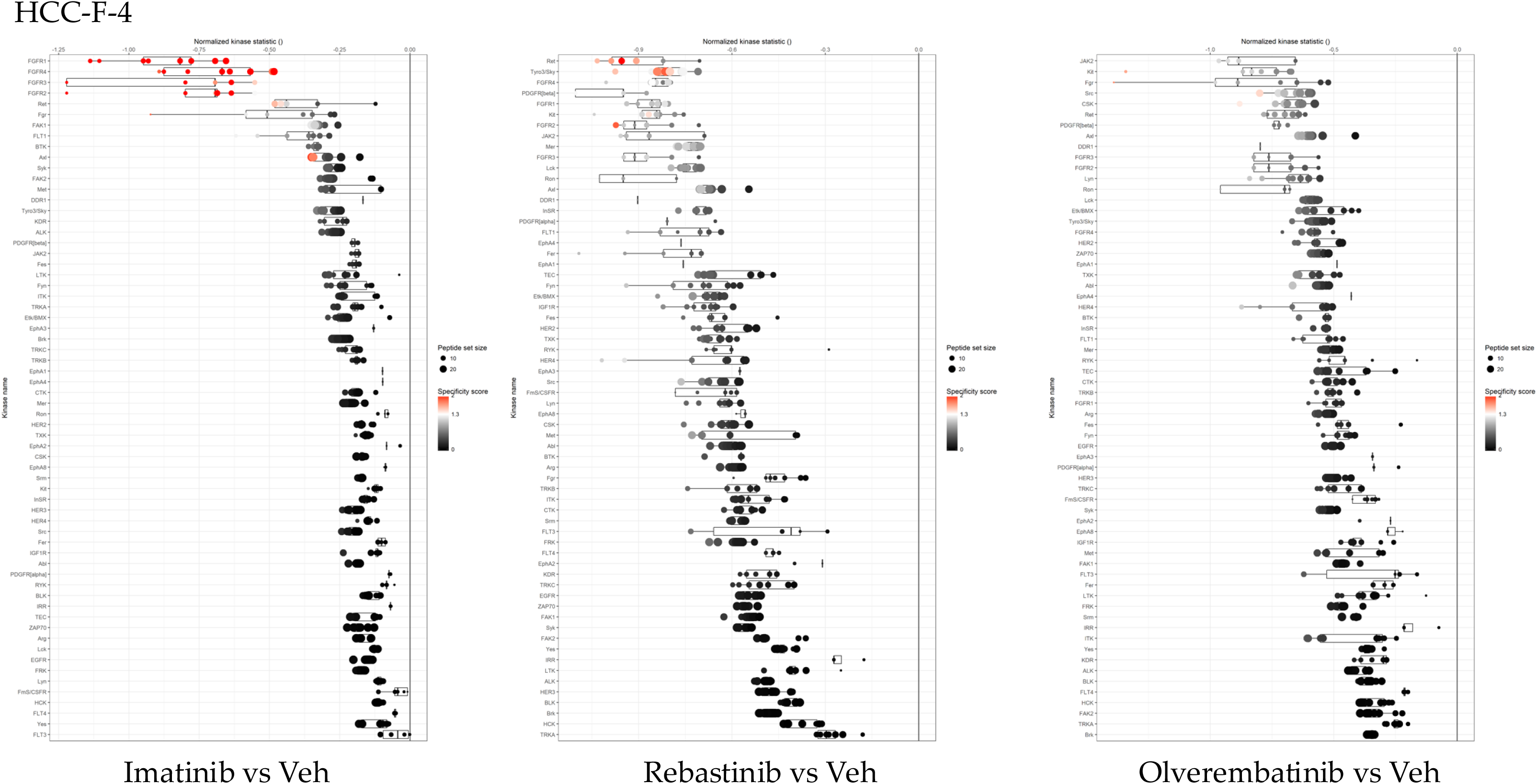
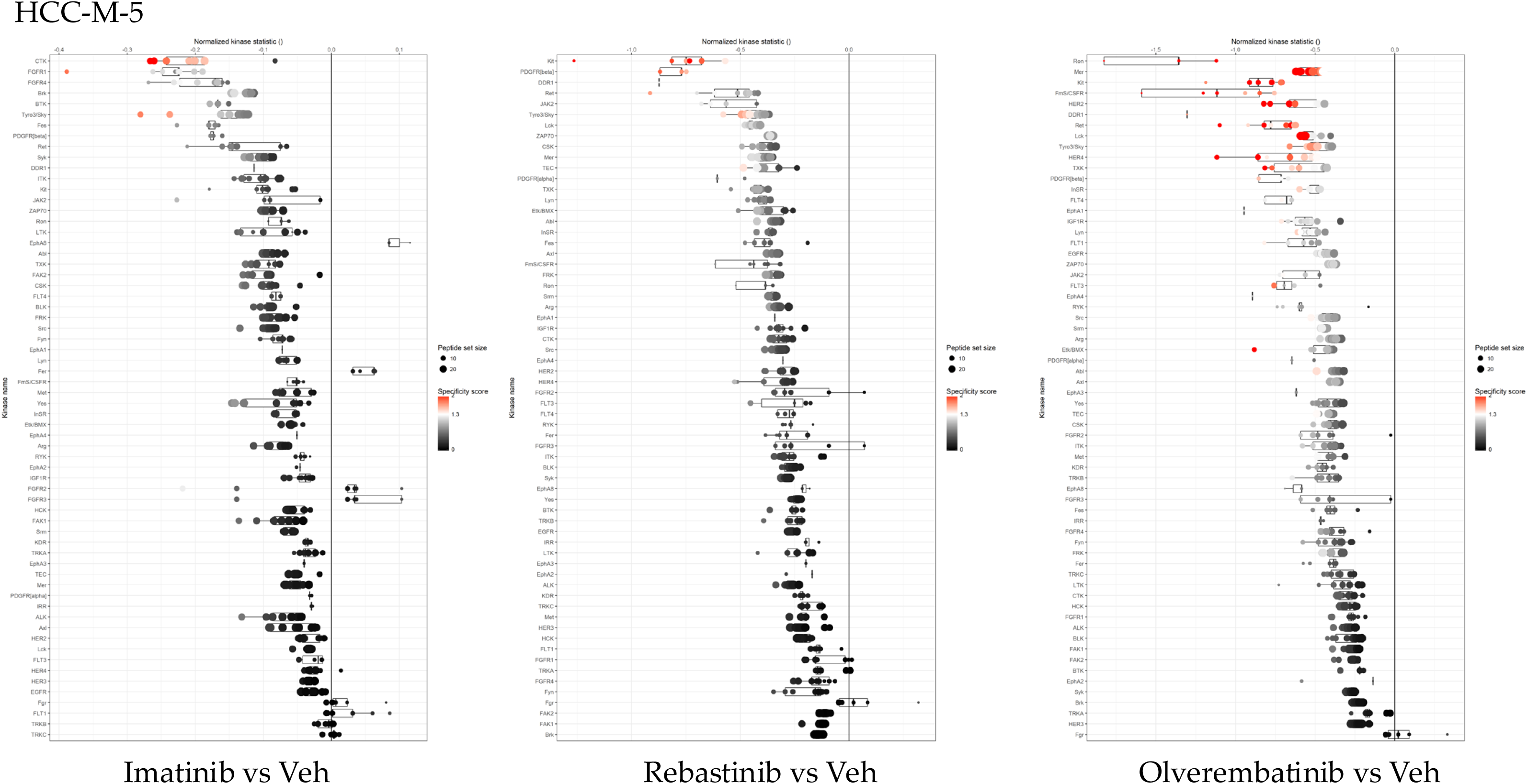
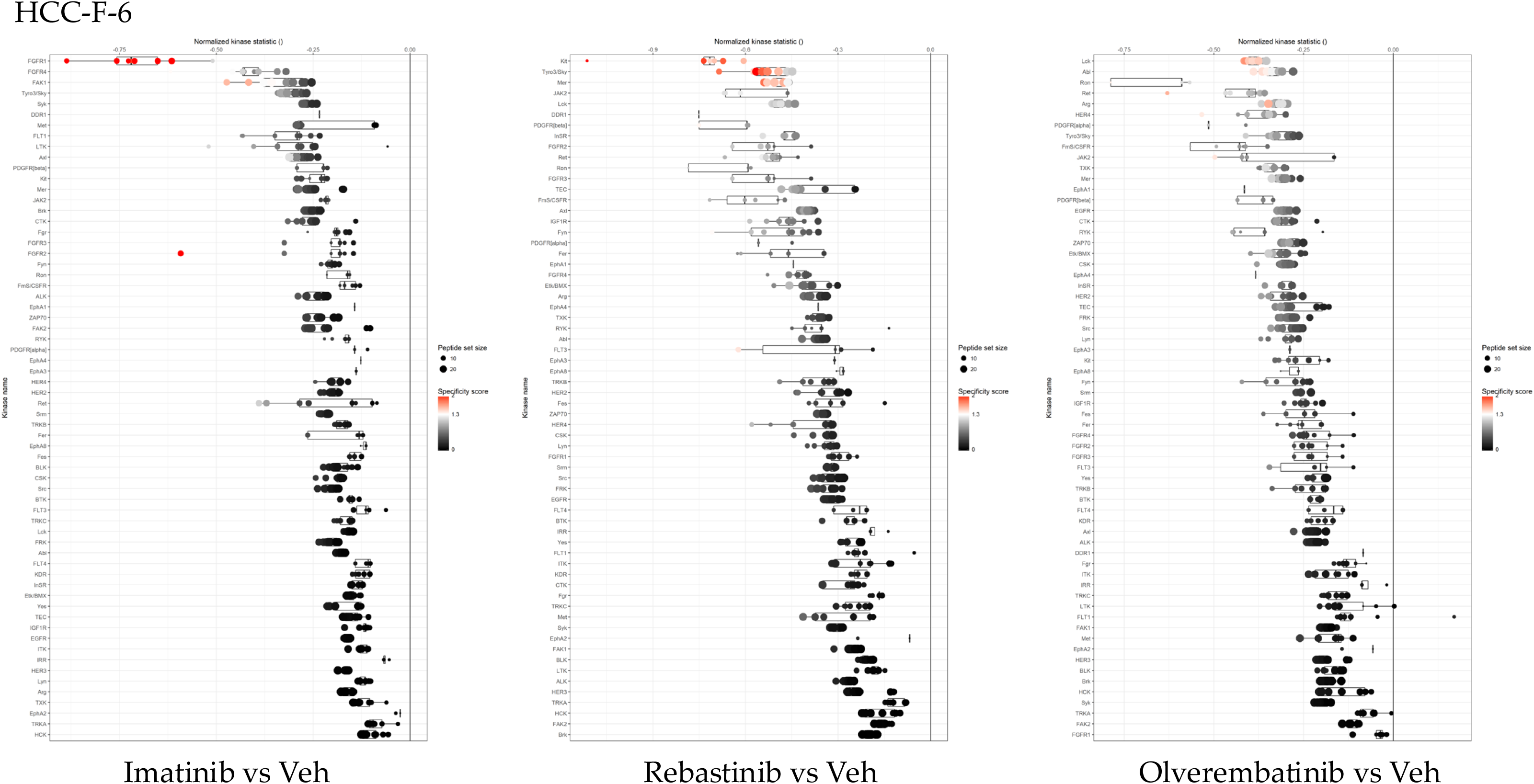
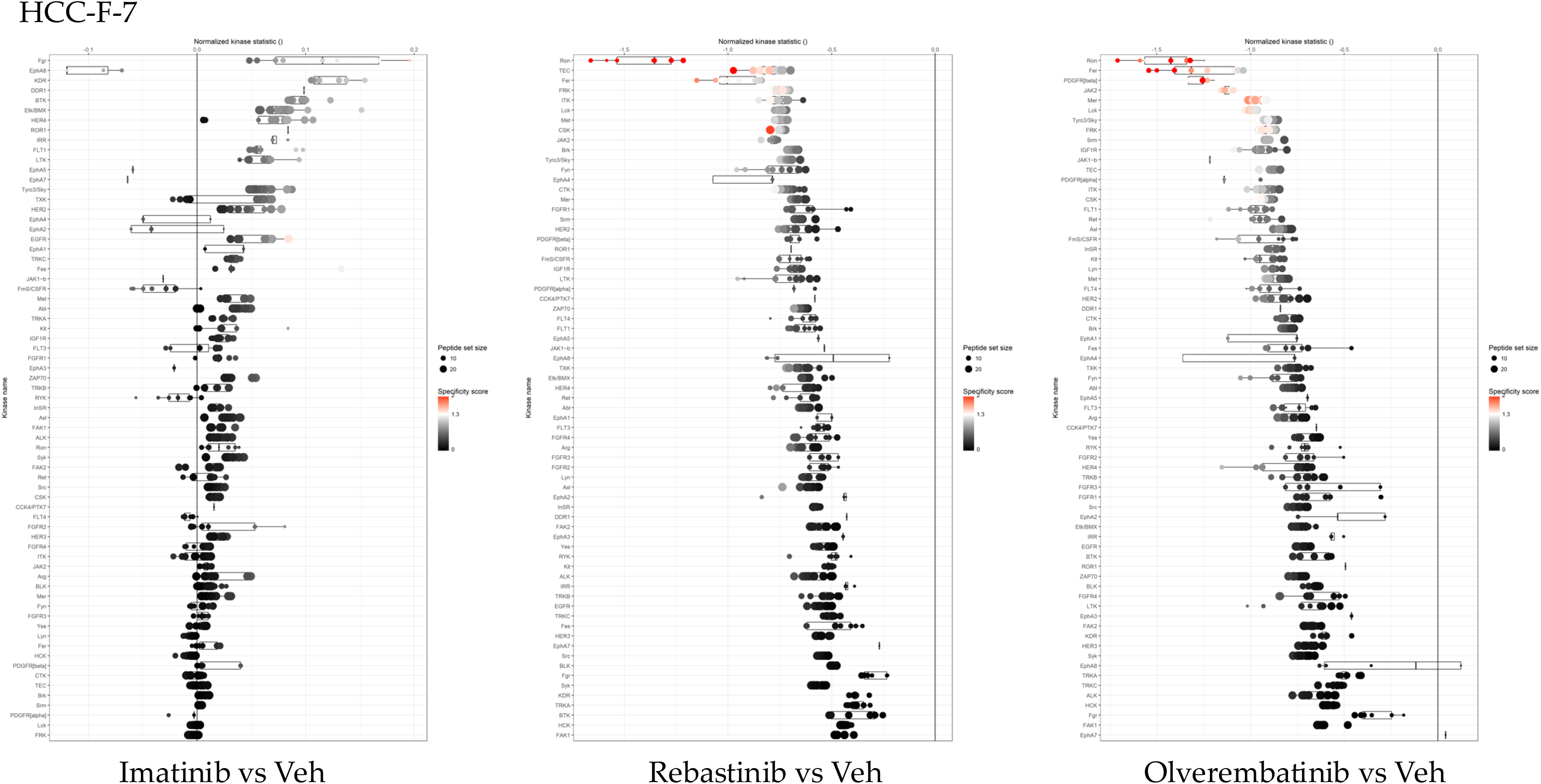
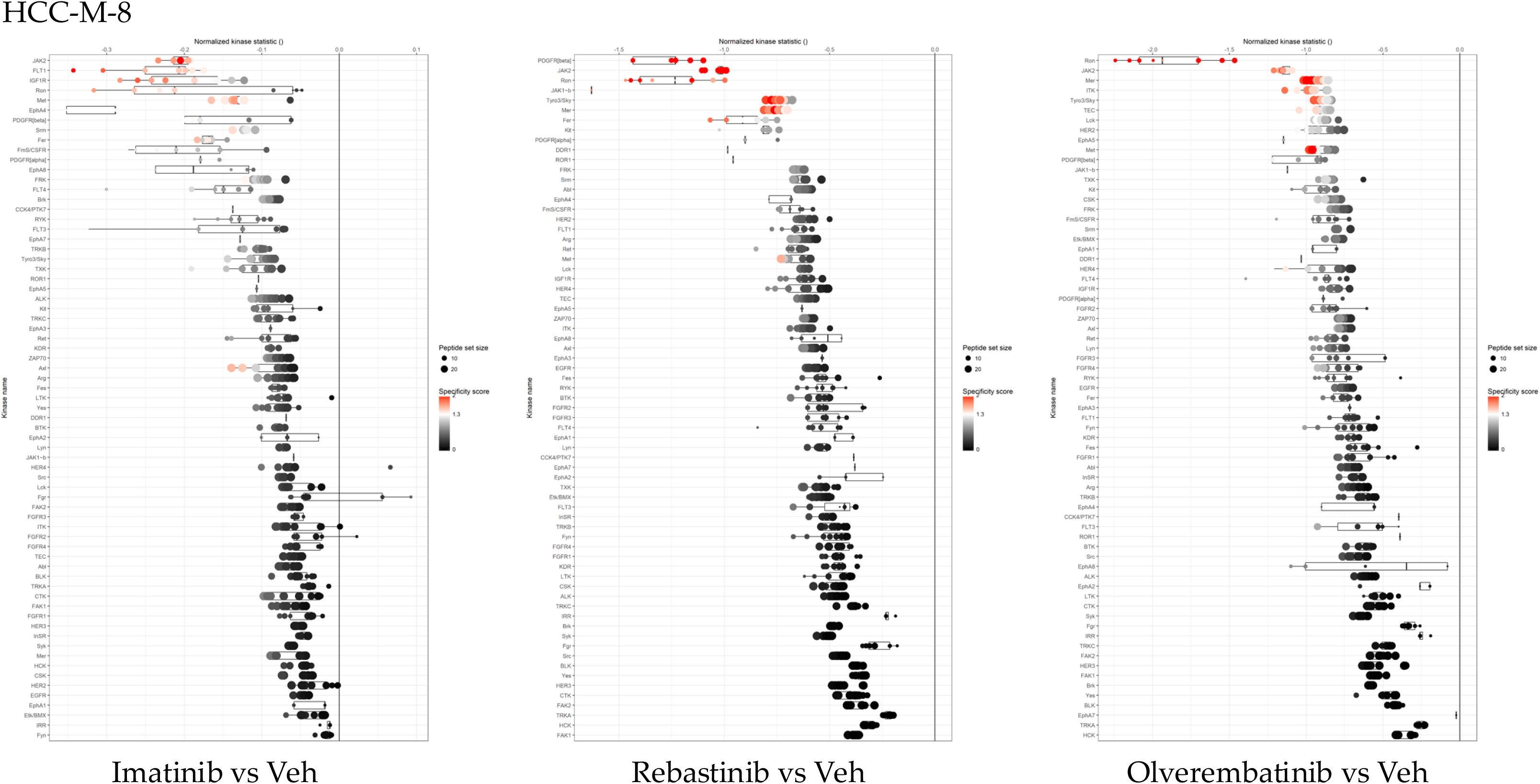
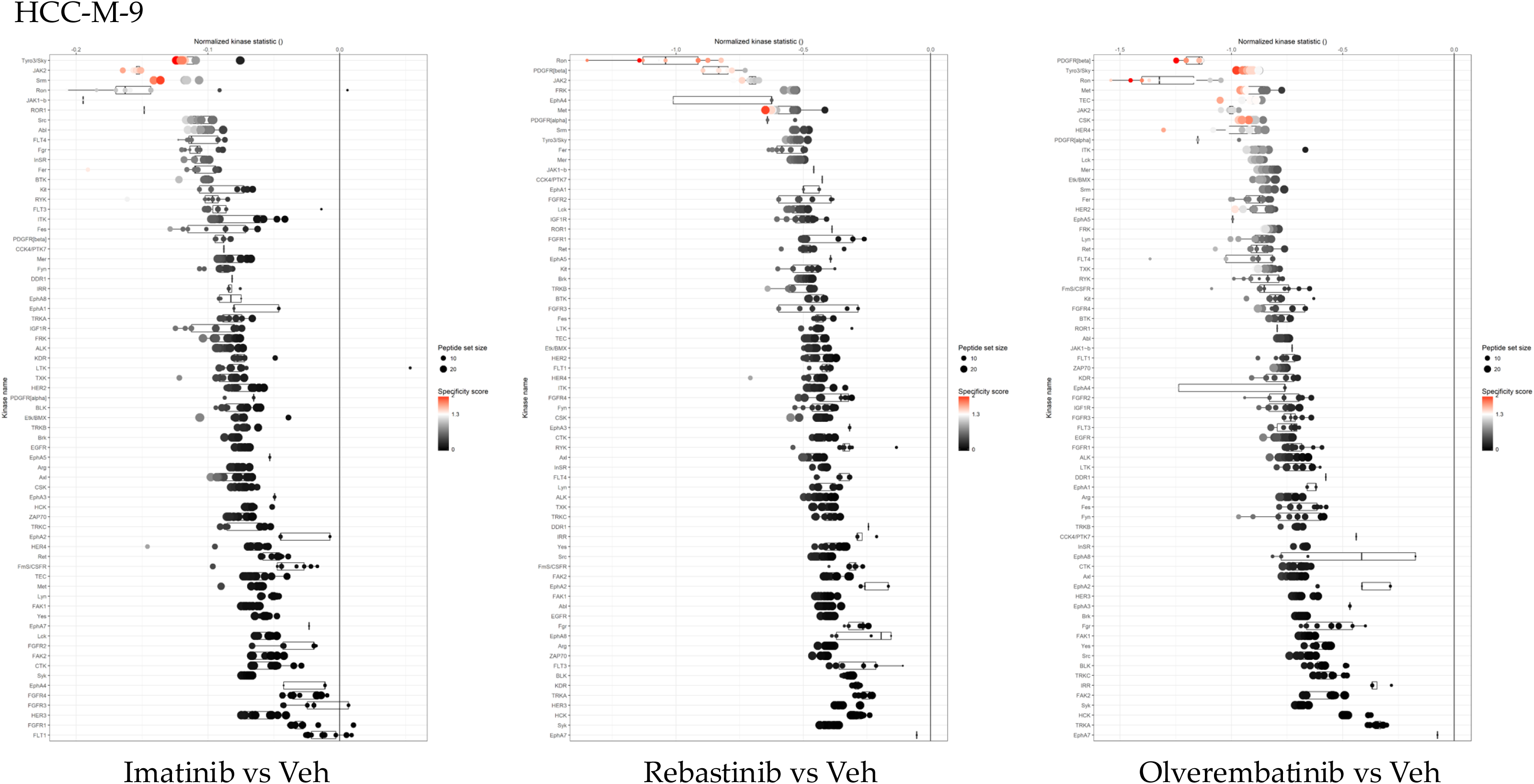

## Appendix B: Individual HCC Samples with ABL Inhibitors

### STK PamChip runs

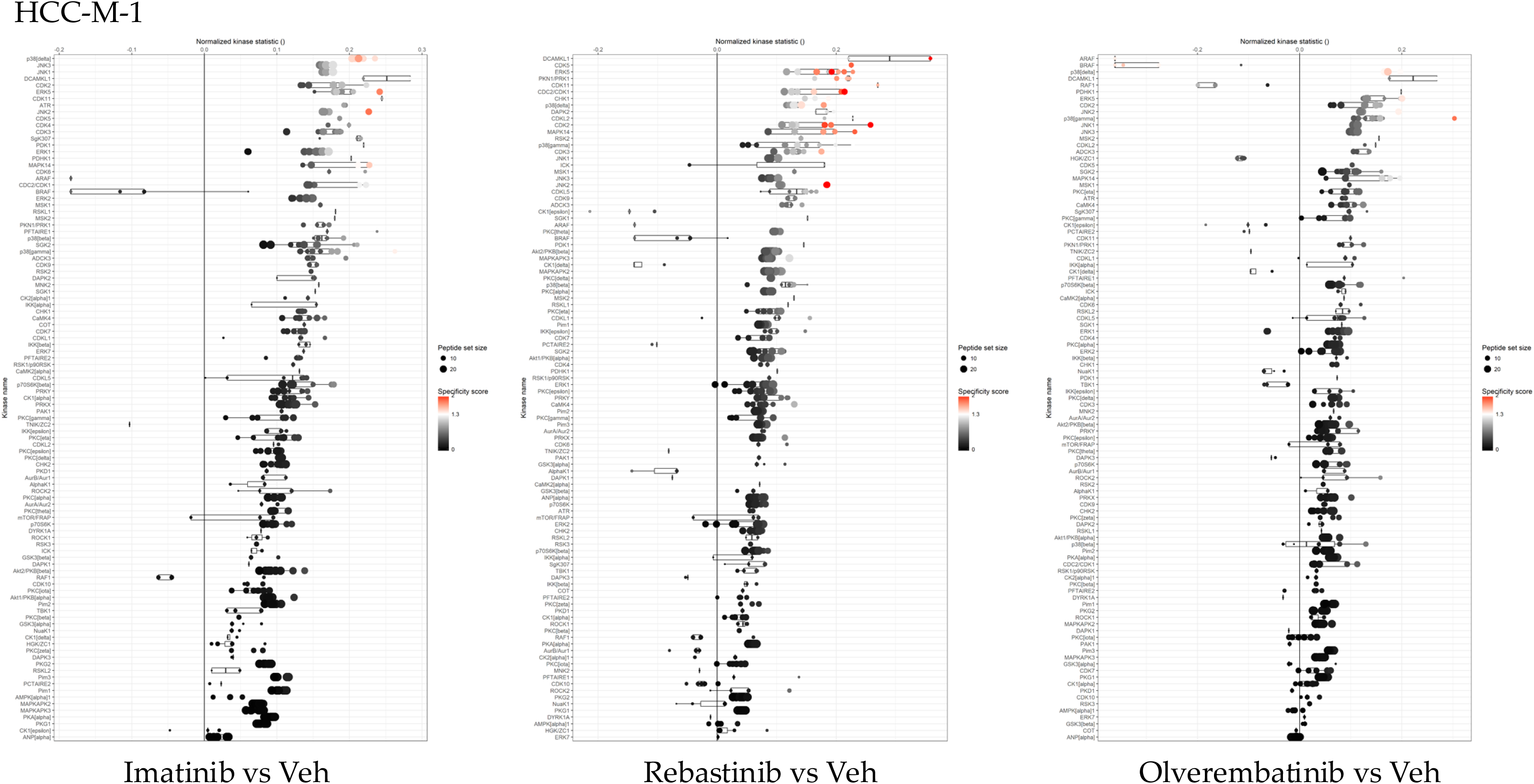
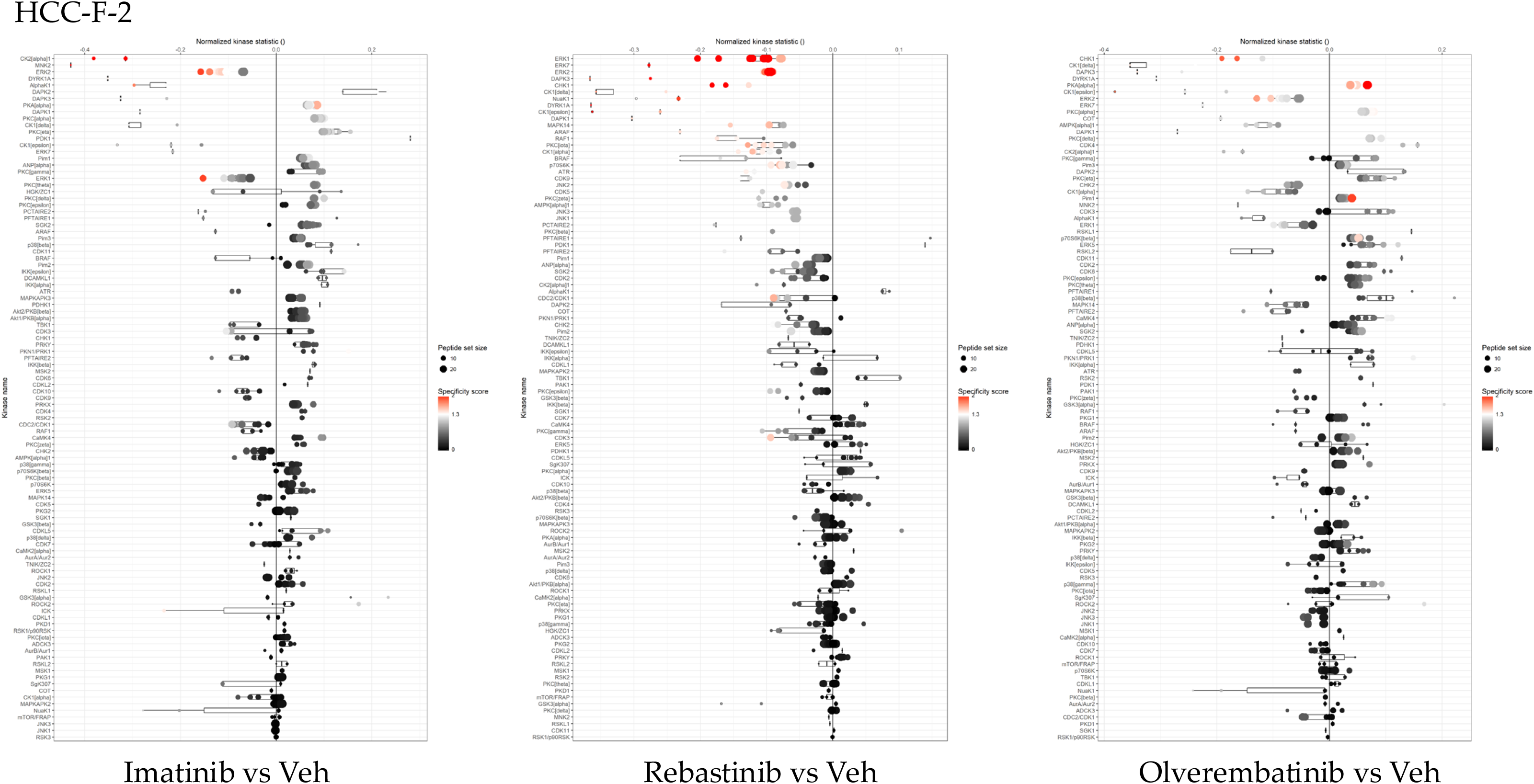
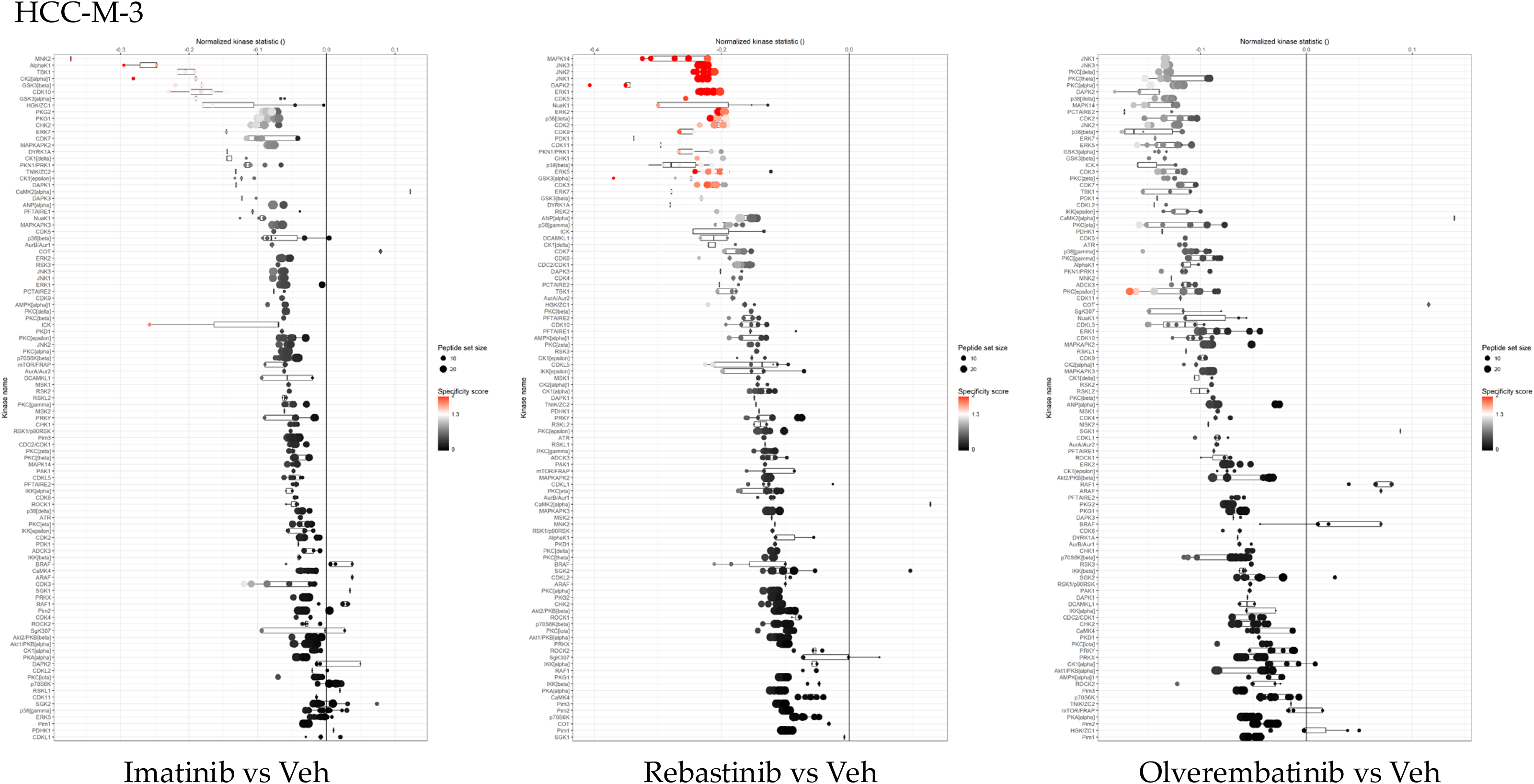
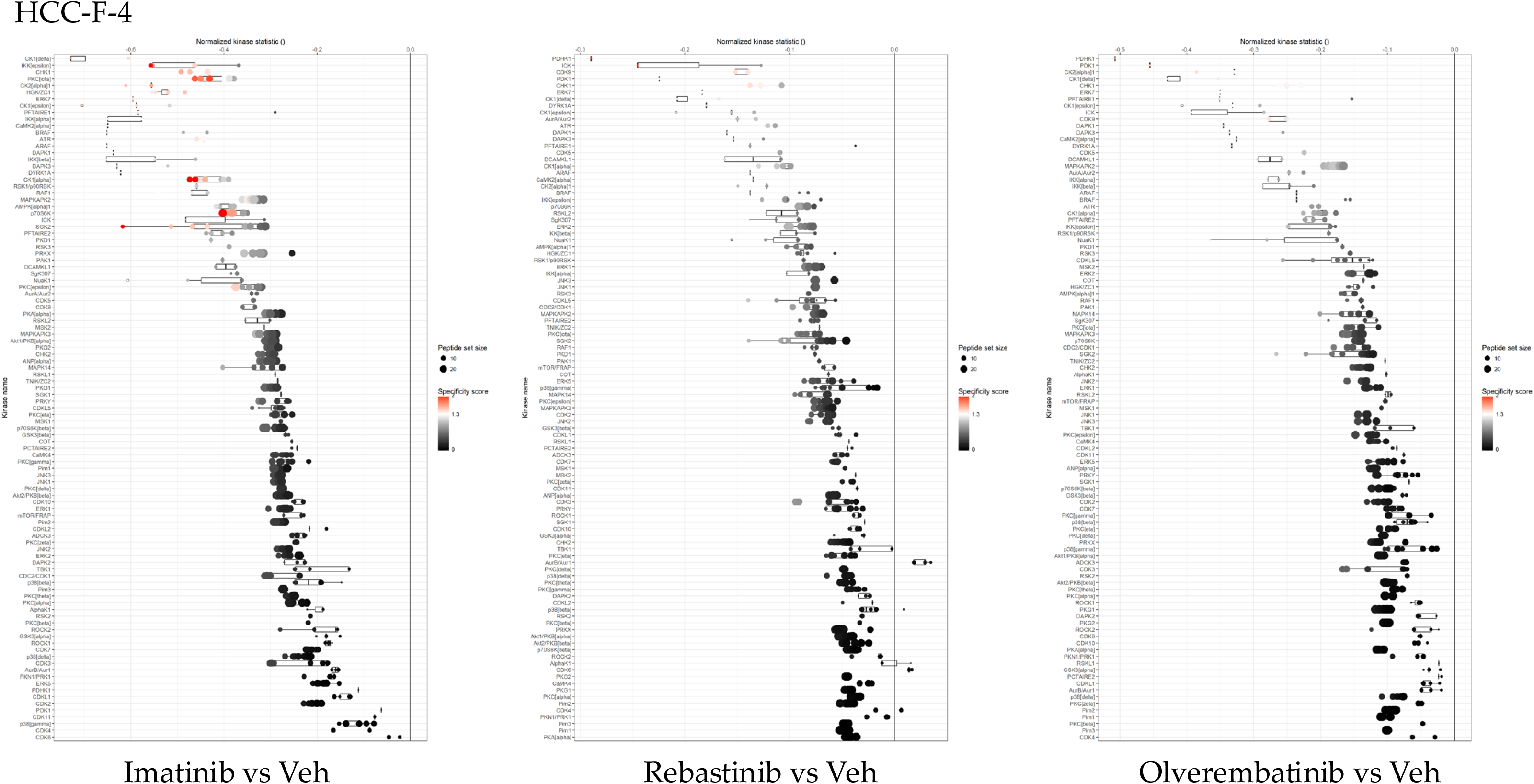
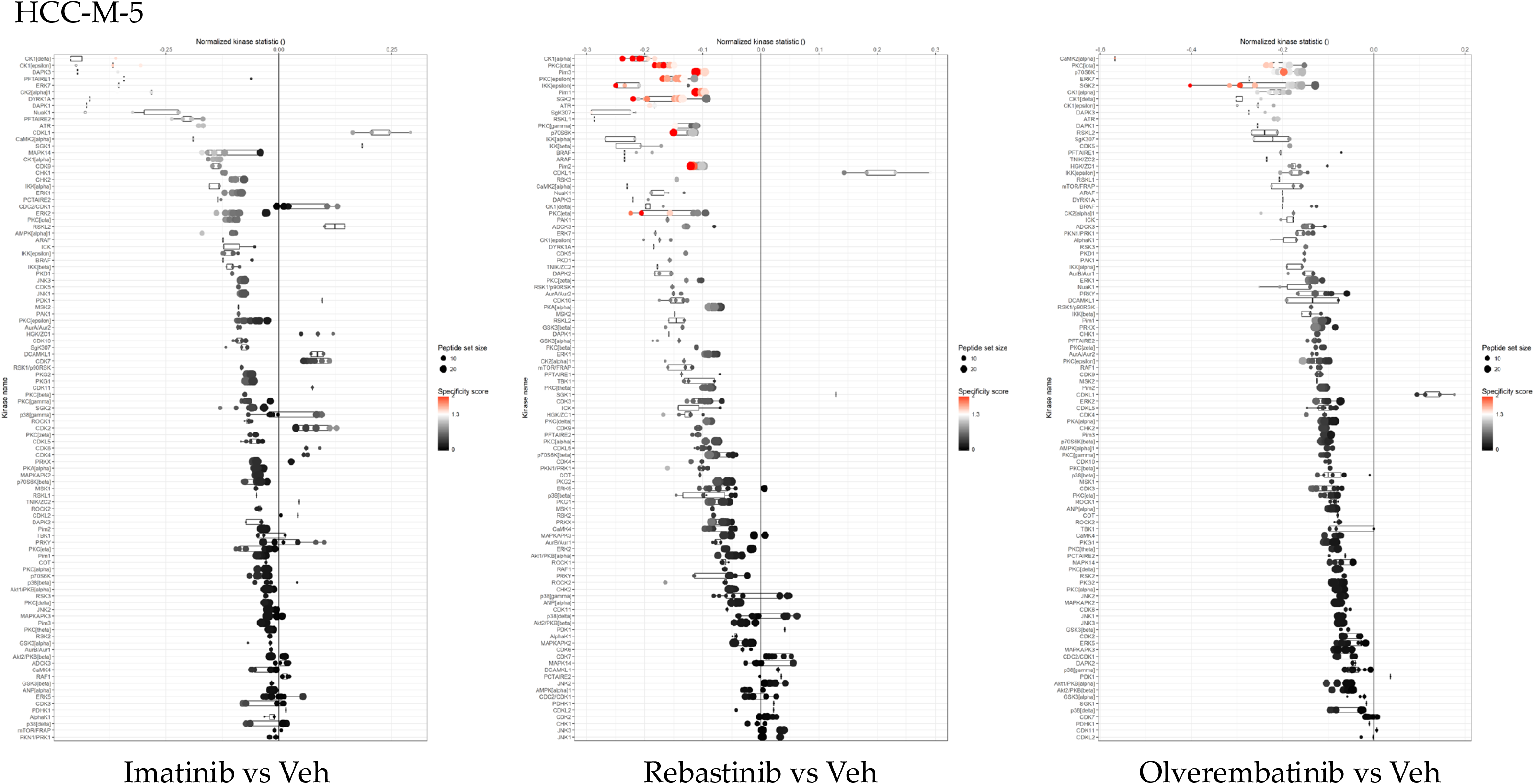
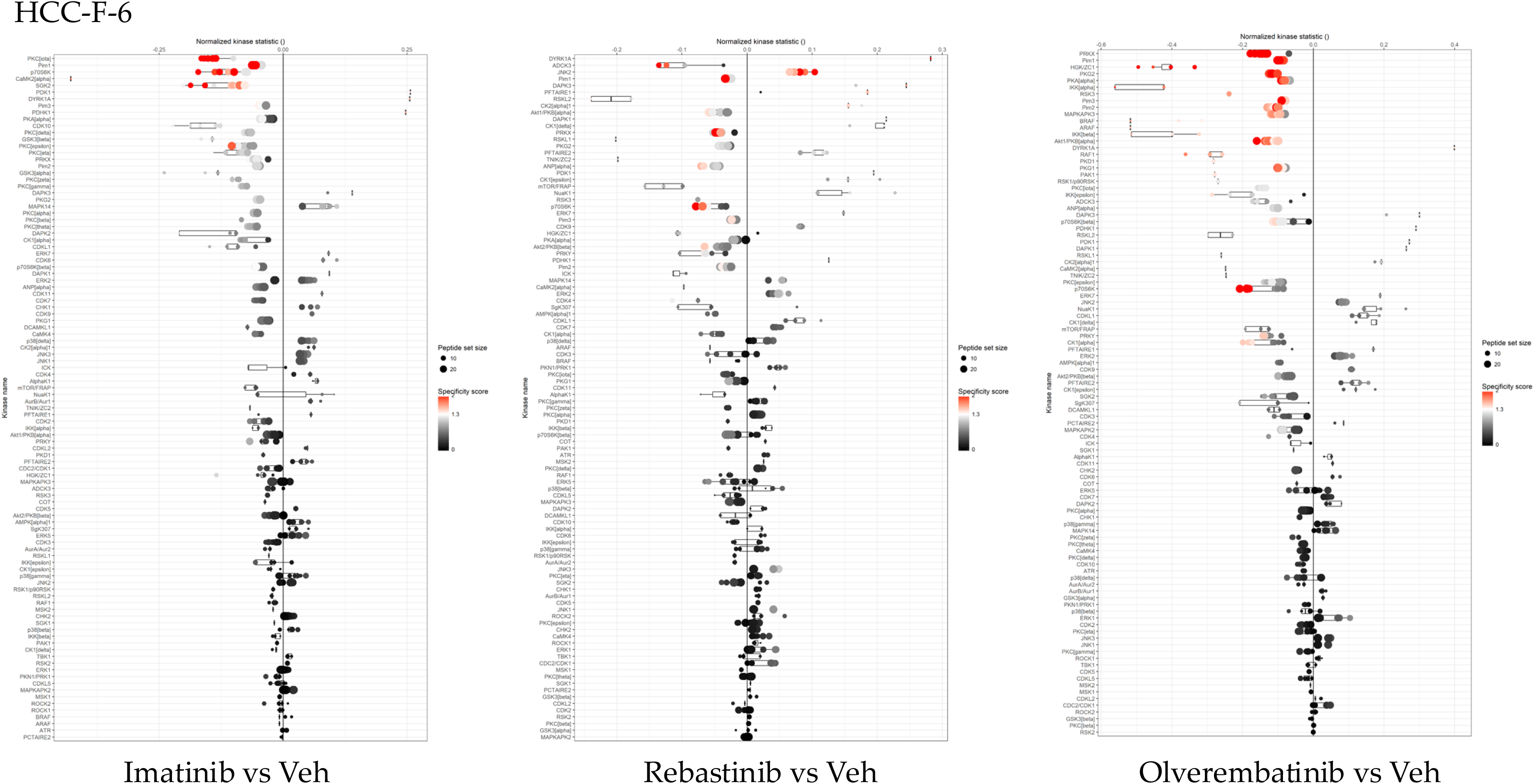
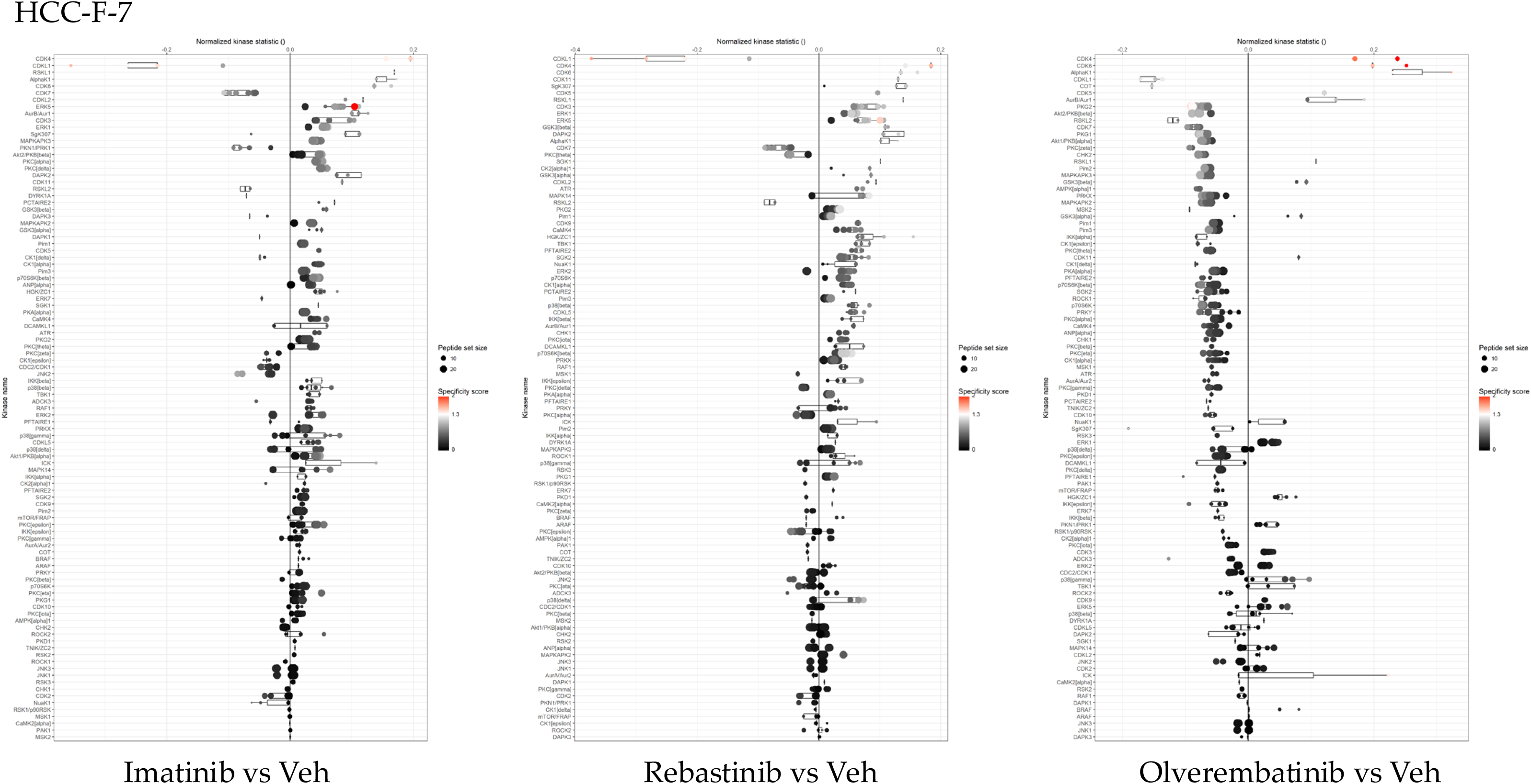
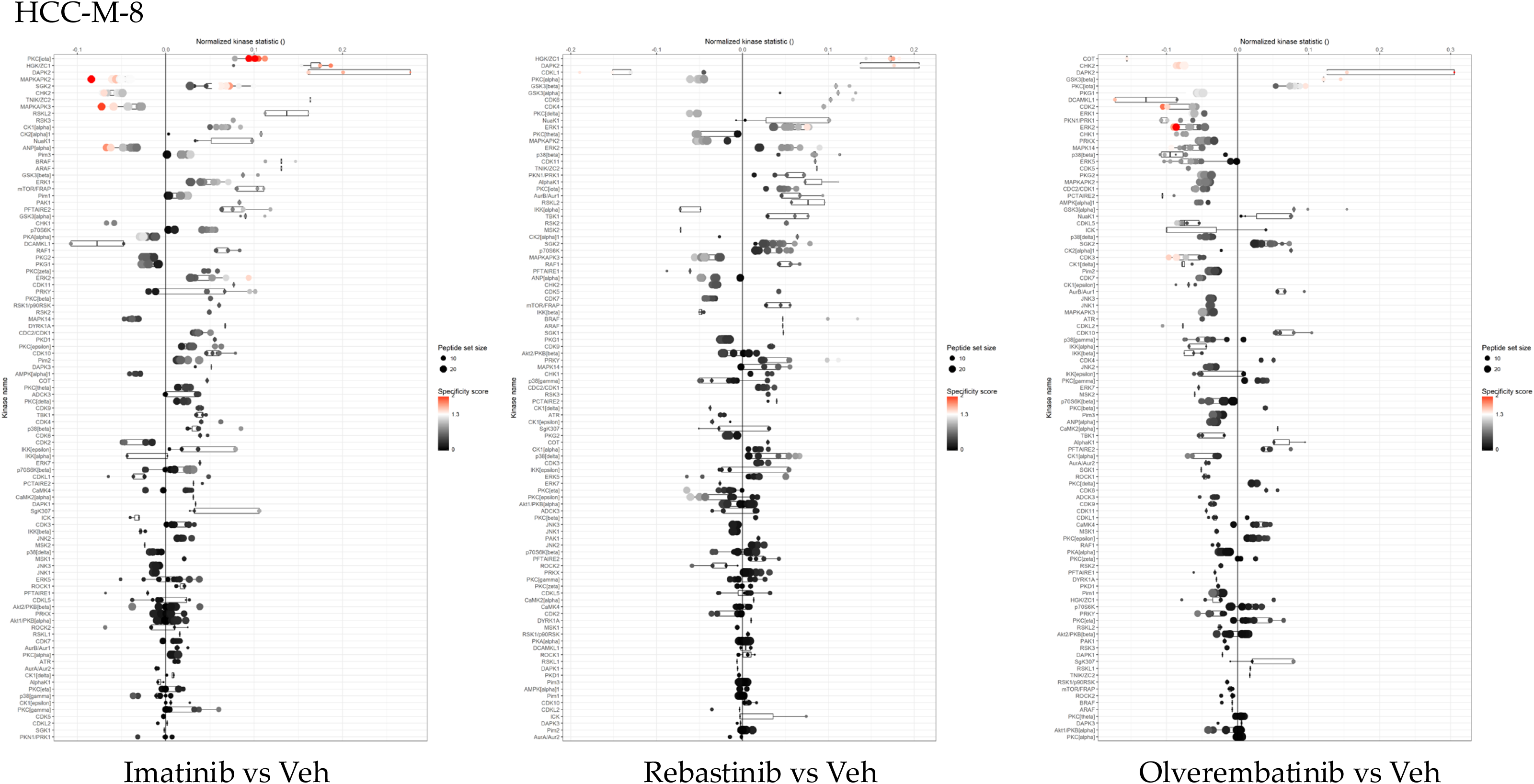
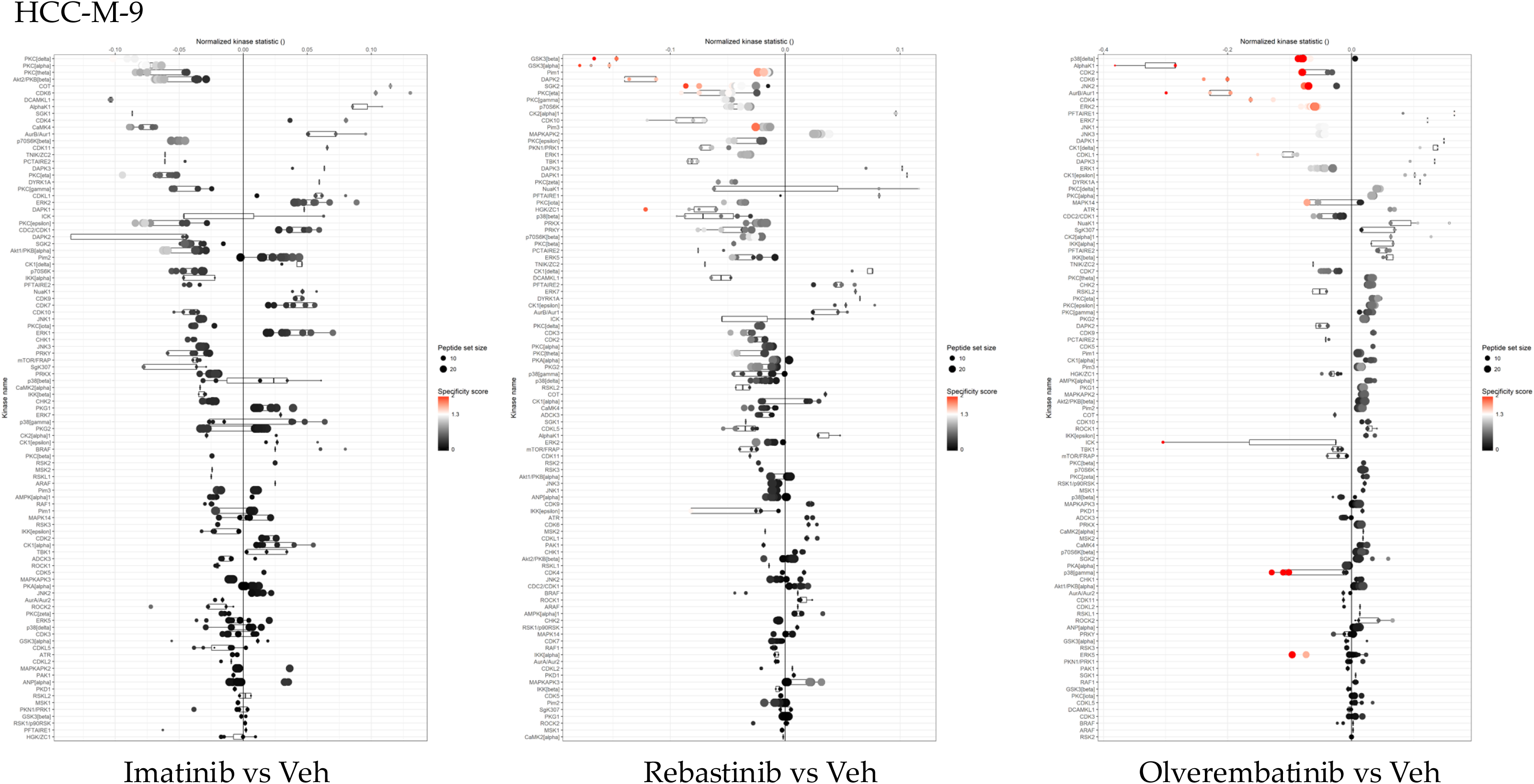

## STAR★METHODS

### KEY RESOURCES TABLE

2 KEY RESOURCES TABLE
| REAGENT or RESOURCE | SOURCE | IDENTIFIER |
| --- | --- | --- |
| Biological samples |  |  |
| Human Liver Biopsies of HCC Tumor and Normal Adjacent Tissue | University of Kentucky Markey Cancer Center Biospecimen Procurement & Translational Pathology Shared Resource Center | https://ukhealthcare.uky.edu/markey-cancer-center/research/srf/bptp |
| Chemicals, peptides, and recombinant proteins |  |  |
| DMEM/F-12 | Gibco | Cat#12500062 |
| DMEM | Gibco | Cat#10566016 |
| William's Medium | Gibco | Cat#32551020 |
| Fetal Bovine Serum | Alkali Scientific | Cat#FB-72 |
| Anti-Anti | Gibco | Cat#15240062 |
| Hydrocortisone | Sigma-Aldrich | Cat#H0888 |
| Insulin Human | Sigma-Aldrich | Cat#I2643 |
| M-PER | Thermo Fisher Scientific | Cat#78501 |
| Protease Inhibitor Cocktail | Sigma-Aldrich | Cat#P2714 |
| Halt Phosphatase Inhibitor | Thermo Fisher Scientific | Cat#78420 |
| Dimethyl Sulfoxide (DMSO) | Sigma-Aldrich | Cat#472301 |
| Imatinib | Selleck Chemicals | Cat#S2475 |
| Rebastinib | Selleck Chemicals | Cat#S2634 |
| Olverembatinib | Selleck Chemicals | Cat#S7194 |
| QIAzol Lysis Reagent | Qiagen | Cat#79306 |
| Chloroform | Sigma-Aldrich | Cat#320331 |
| Ethanol 200 Proof | KOPTEC | DSP-MD-43 |
| TrueAMP SYBR Green qPCR SuperMix | Alkali Scientific | Cat#75013 |
| Critical commercial assays |  |  |
| Pierce Microplate BCA Protein Assay Kit | Thermo Fisher Scientific | Cat#23252 |
| Tyrosine Kinase PamChip-4 | PamGene | Cat#32516 |
| PTK Reagent Kit | PamGene | Cat#32112.5 |
| Serine/Threonine Kinase PamChip-4 | PamGene | Cat#32501 |
| STK Reagent Kit | PamGene | Cat#32201.3 |
| RNeasy Mini Kit | Qiagen | Cat#74106 |
| High Capacity cDNA Reverse Transcription Kit | Applied Biosystems | Cat#4368814 |
| Deposited data |  |  |
| Full Kinome Reports and Raw Kinome Files | This Paper | Figshare: DOI |
| Experimental models: Cell lines |  |  |
| HepG2 | Samir Softic | N/A |
| HLE | Robert Helsley | N/A |
| Huh-7 | Sadeesh Ramakrishnan | N/A |
| Hep3B2 | ATCC | LOT: 63194732 |
| HepaRG | Thermo Fisher Scientific | Cat#HPRGC10 |
| Oligonucleotides |  |  |
| Primers for RT-PCR, See Table S4 | This Paper | N/A |
| Software and algorithms |  |  |
| Evolve 3 | PamGene | https://pamgene.com |
| BioNavigator | PamGene | https://pamgene.com |
| RStudio | Posit Software | https://posit.co |
| KRSA | DePasquale et al. <sup>1</sup> | https://github.com/CogDisResLab/KRSA |
| CORAL | Metz et al. <sup>2</sup> | http://phanstiel-lab.med.unc.edu/CORAL/ |
| Prism 10.4 | GraphPad Software | https://www.graphpad.com |
| Other |  |  |
| PamStation 12 | PamGene | https://pamgene.com |

### EXPERIMENTAL MODEL AND STUDY PARTICIPANT DETAILS

#### Hepatocellular Carcinoma Samples

Human liver biopsies of hepatocellular carcinoma and normal adjacent tissue were received from the Biospecimen Procurement & Translational Pathology Shared Resource Facility (BPTP SRF) of the University of Kentucky Markey Cancer Center (P30CA177558). The samples were deidentified to the research group and designated non-human research with IRB exemption. The H&E images in Figure 1 were provided by BPTP SRF. The clinical characteristics of the study population are in Table S1.

#### Human cell lines

HepG2 and Huh-7 cells were cultured in DMEM/F12 medium (Gibco, 12500062) supplemented with 10% fetal bovine serum (FBS, Alkali Scientific, FB-72) and 1% Anti-Anti (AA, Gibco, 15240062). HLE and Hep3B2 cells were cultured in DMEM (Gibco, 10566016) supplemented with 10% FBS and 1% AA. HepaRG cells were cultured in Willams medium (Gibco, 32551020) supplemented with 10% FBS, 1% AA, 24.7 μg/mL hydrocortisone (Sigma-Aldrich, H0888), and 5 μg/mL insulin (Sigma-Aldrich, I2643). All cells were grown at 37°C with 5% CO2 in a humidified incubator.

### METHOD DETAILS

#### PamGene PamStation Kinome Analysis

Kinase activity was quantified in the HCC and normal adjacent tissue samples using the PamGene PamStation 12 instrument, as previously described ^3–7^. Samples were cut and pooled within their respective group and lysed using Qiagen TissueLyser and M-PER protein extraction reagent (Thermo Fisher Scientific, 78501) supplemented with protease inhibitor (Sigma-Aldrich, P2714) and Halt phosphatase inhibitor (Thermo Fisher Scientific, 78420). The samples were centrifuged, and the supernatant was transferred to a new tube. Protein concentration was determined by Pierce Microplate BCA Protein Assay Kit (Thermo Fisher Scientific, 23252). The samples were diluted to a 2.5 μg/μL concentration in lysis buffer. Per array for STK PamChip (PamGene, 32501), 1 μg of protein was used, and 5 μg was used for PTK PamChip (PamGene, 32516). The lysates were added to the PamChip in the presence of 4 mM adenosine triphosphate (ATP) and fluorescently labeled antibodies (PamGene, 32112.5 PTK, 32201.3 STK) to detect the phosphorylation of 196 (PTK) or 144 (STK) peptide substrates per array. The samples were run in technical triplicates using 3 PamChips for each PTK and STK. Evolve 3 (PamGene) software uses a charge-coupled device (CCD) camera and light-emitting diode (LED) imaging system to record relative phosphorylation levels of each unique consensus phosphopeptide sequence every 5 min for 60 min as measured by peptide signal intensities recorded across 10, 20, 50, and 100 millisecond exposure times over 94 cycles for PTK and 124 cycles for STK. Raw imaging data were exported for further data analysis, and kinase mapping was described further in the BioNavigator Bioinformatics Analysis and Kinome Random Sampling Analysis (K.R.S.A) section.

#### Clinical Trial in a PamChip

Protein lysate from the men and women HCC tumor region prepared above from the kinome analysis were used to perform the clinical trial in a PamChip. A similar protein lysis method as described above was used for the individual HCC samples. For the HepG2, Huh-7, HLE, Hep3B2, and HepaRG validation runs protein was harvested 24 hours after a media change with normal culture medium described above. The cells were scraped, and protein was extracted using M-PER supplemented with protease and phosphatase inhibitors as described above and cell pellets were homogenized using a pellet pestle. A similar protocol as described above for the PamGene PamStation Kinome Analysis was used with two modifications. The same amount of protein lysate described above was used. The protein lysates and run basic mix were incubated with either vehicle (DMSO, Sigma-Aldrich, 472301), Imatinib (1 μM) (Selleckchem, S2475), Rebastinib (100 nM) (Selleckchem, S2634), or Olverembatinib (100 nM) (Selleckchem, S7194) for 30 minutes at room temperature while rocking prior to the addition of the ATP and fluorescently-label antibody. The concentrations were determined using the IC50 concentrations published on Selleckchem.com. The same final concentration of DMSO (2%) was used for all treatments. The concentration of ATP was lowered to 1 mM to ensure the concentration of ATP would not out- compete the inhibitors used. The run conditions were the same as above. The raw images were exported for quantification and further analysis described below.

#### BioNavigator Bioinformatics Analysis

The images of the arrays taken during the runs were analyzed using BioNavigator (PamGene). The fold change (FC) for each phosphopeptide was calculated using the signal ratios and averaged across the three PamChips. The minimum threshold cutoffs were determined from previous literature ^3–7^. These thresholds require differential phosphopeptide signals greater than or equal to 30% (FC ≥ 1.30 or FC ≤ 0.70) for differential phosphorylation to be considered. Linear regression slopes provide phosphorylation intensity signals over time that are used to compare groups (e.g., experimental vs. control). Data was exported from BioNavigator after calculation of signal intensity for further data analysis. Upstream kinase identification was performed using BioNavigator Upstream Kinase Analysis (UKA) (PamGene). The UKA analysis identifies the kinases most likely responsible for phosphorylation events occurring on the PamChip over time. The log transformed signal intensity for the substrates were determined, compared to the normal adjacent tissue or vehicle, and used for the PerMed plots. The kinase activity plots were made using Prism (GraphPad). For the kinome analysis of HCC tissue, the data was analyzed by comparing the HCC tumor region to the normal adjacent tissue for each sex. For the Clinical Trial in a PamChip, the data was analyzed by comparing each inhibitor to the vehicle for each sex for the liver samples, individual HCC samples, and cell lines.

#### Kinome Random Sampling Analysis (K.R.S.A.)

The signal intensity and saturation data were exported into an excel file for further analysis using the Kinome Random Sampling Analysis (K.R.S.A) package on RStudio. Undetectable and/or nonlinear (R^2^ < 0.80) phosphopeptides are excluded from subsequent analyses. The data was analyzed by comparing the HCC tumor region to the normal adjacent tissue for the liver samples. For the Clinical Trial in a PamChip, the data was analyzed by comparing each inhibitor to the vehicle for each sex for the liver samples, individual HCC samples, and cell lines. Full kinome reports and raw data files are available on Figshare (DOI TBD).

#### CORAL Kinome Phyla Trees

The kinome phyla tree was made using CORAL^2^. The phyla tree values were generated and exported from BioNavigator. Each node represents a kinase, and kinases are grouped by family. Only kinases measured in our analysis are included on the tree. The node size for each kinase refers to the mean final score for direction of kinase activity. The node color represents the median kinase statistic, which reflects the magnitude of kinase activity changes. Value ranges for node color and node size are based on the ranges of all values in the analysis and are consistent for comparison with the other phyla trees in the figure.

#### RNA Extraction and Quantitative Real-Time PCR

RNA was extracted from the HCC and normal adjacent tissues for gene expression quantification using RT-PCR. The tissues were lysed in QIAzol (Qiagen, 79306) and homogenized using Qiagen TissueLyser. The RNA was extraction using phenol:chloroform separation by the addition of chloroform (Sigma-Aldrich, 320331) to the QIAzol. The upper supernatant was transferred to a new tube containing ethanol (KOPTEC, DSP-MD-43). The total RNA was purified using a RNeasy Mini Kit (Qiagen, 74106) following manufacture provided instructions. The total RNA was quantified using a NanoDrop 2000 spectrophotometer (Thermo Fisher Scientific). Using total RNA, cDNA was synthesized using High Capacity cDNA Reverse Transcription Kit (Applied Biosystems, 4368814). RT-PCR was using to amplify and quantify the cDNA gene expression using specific primers (see Table S4) and TrueAMP SYBR Green qPCR SuperMix (Alkali Scientific, 75013). The thermocycling conditions consisted of: 95°C for 10 minutes, 60 cycles of 95°C for 15 seconds, 60°C for 30 seconds and 72°C for 0-30 seconds based on primer product size and finished with a melt curve with temperatures ranging from 60-95°C to determine primer specificity. Normalization was performed in a separate reaction to RPL41. The RT-PCR graphs were made using Prism (GraphPad).

### QUANTIFICATION AND STATISTICAL ANALYSIS

All RT-PCR graphs contain an individual point per sample (n # shown in Table S1) on the graph. The kinase activity graphs have a gray diamond for each kinase target substrate measured. The PamChip images are a representative image of one out of the three chips ran in the run. RT-PCR statistics were calculated using a Wilcoxon Test on Prism. p values < 0.05 were considered statistically significant.

